# Phylogenetic analysis of SARS-CoV-2 in the Boston area highlights the role of recurrent importation and superspreading events

**DOI:** 10.1101/2020.08.23.20178236

**Authors:** Jacob E. Lemieux, Katherine J. Siddle, Bennett M. Shaw, Christine Loreth, Stephen F. Schaffner, Adrianne Gladden-Young, Gordon Adams, Timelia Fink, Christopher H. Tomkins-Tinch, Lydia A. Krasilnikova, Katherine C. DeRuff, Melissa Rudy, Matthew R. Bauer, Kim A. Lagerborg, Erica Normandin, Sinead B. Chapman, Steven K. Reilly, Melis N. Anahtar, Aaron E. Lin, Amber Carter, Cameron Myhrvold, Molly E. Kemball, Sushma Chaluvadi, Caroline Cusick, Katelyn Flowers, Anna Neumann, Felecia Cerrato, Maha Farhat, Damien Slater, Jason B. Harris, John Branda, David Hooper, Jessie M. Gaeta, Travis P. Baggett, James O’Connell, Andreas Gnirke, Tami D. Lieberman, Anthony Philippakis, Meagan Burns, Catherine M. Brown, Jeremy Luban, Edward T. Ryan, Sarah E. Turbett, Regina C. LaRocque, William P. Hanage, Glen R. Gallagher, Lawrence C. Madoff, Sandra Smole, Virginia M. Pierce, Eric Rosenberg, Pardis C. Sabeti, Daniel J. Park, Bronwyn L. Maclnnis

## Abstract

SARS-CoV-2 has caused a severe, ongoing outbreak of COVID-19 in Massachusetts with 111,070 confirmed cases and 8,433 deaths as of August 1, 2020. To investigate the introduction, spread, and epidemiology of COVID-19 in the Boston area, we sequenced and analyzed 772 complete SARS-CoV-2 genomes from the region, including nearly all confirmed cases within the first week of the epidemic and hundreds of cases from major outbreaks at a conference, a nursing facility, and among homeless shelter guests and staff. The data reveal over 80 introductions into the Boston area, predominantly from elsewhere in the United States and Europe. We studied two superspreading events covered by the data, events that led to very different outcomes because of the timing and populations involved. One produced rapid spread in a vulnerable population but little onward transmission, while the other was a major contributor to sustained community transmission, including outbreaks in homeless populations, and was exported to several other domestic and international sites. The same two events differed significantly in the number of new mutations seen, raising the possibility that SARS-CoV-2 superspreading might encompass disparate transmission dynamics. Our results highlight the failure of measures to prevent importation into MA early in the outbreak, underscore the role of superspreading in amplifying an outbreak in a major urban area, and lay a foundation for contact tracing informed by genetic data.

## Main Text

SARS-CoV-2 has now caused over 22 million infections and over 775,000 deaths worldwide (*1*) in one of the worst public health crises of the past century. The early impact of the COVID-19 pandemic has been particularly severe in the state of Massachusetts (MA) in the northeastern United States (US). The first case in the state was confirmed on February 1, 2020 (*2*); case counts rapidly accelerated beginning in March and peaked in the third week in April. The Boston area, home to 70% of the population of MA, accounted for 79% of COVID-19 cases and 76% of COVID-19 deaths in the state to this point (*3*). COVID-19 has disproportionately affected vulnerable populations, particularly residents and staff in congregate living environments (*4*) and racial and ethnic minorities (*5*, *6*). In MA, residents and healthcare workers in long-term care facilities accounted for 22% of all confirmed cases of COVID-19 and 64% of all reported deaths through August 1, 2020 (*7*).

COVID-19, like previous coronavirus outbreaks (*8*, *9*), has been marked by the prominence of superspreading events (*10*, *11*), in which one individual infects an unusually large number of secondary cases. (For this study, we define a superspreading event as the transmission of at least 8 secondary infections from a single source, corresponding to the 99th percentile (*12*) for an R_eff_ of 2.5.) More broadly, a great deal of SARS-CoV-2 transmission has occurred in clusters of cases linked to events and gatherings, including on cruise ships (*13*), in churches (*14*), and especially in congregate settings such as care homes (*15*), homeless shelters (*16*), and prisons (*17*). However, the evidence indicating that these events drive transmission has been based largely on time-series data showing an increase in cases following them (*18*). Case counts alone have little ability to determine the contribution of any event to overall transmission or to distinguish superspreading from other forms of locally intense transmission. Yet understanding how the virus is actually spreading is critical for prioritizing public health interventions: cluster-based spread may be controlled with more limited restrictions than the population measures required to curb community-based transmission. Genomic data can reveal connections between cases that cannot be detected through conventional epidemiology alone, including direct evidence of superspreading based on shared viral sequences. To gain insight into the introduction and spread of SARS-CoV-2, and to examine the role of putative transmission linked to events and gatherings, we conducted a detailed genomic epidemiology study of the Boston area epidemic.

## Genomic Analysis of SARS-CoV-2 from the Boston Area

We performed viral genome sequencing and phylogenetic analysis of SARS-CoV-2-positive nasopharyngeal (NP) samples collected by the Massachusetts Department of Public Health (MADPH) between January 29, 2020, and April 18, 2020, and by the Massachusetts General Hospital (MGH) between March 4, 2020, and May 9, 2020. Our dataset includes nearly all confirmed early cases of the epidemic in MA through March 8, 2020 (Fig. 1A-B); samples from many of the highest-prevalence communities in and around Boston across the first wave (Fig. 1C), including Chelsea, Revere, and Everett (Fig. 1D-F, Fig. S1); and samples from putative superspreading events involving an international conference and congregate living environments, specifically among homeless shelter guests and staff and within a skilled nursing facility.

**Fig 1.**
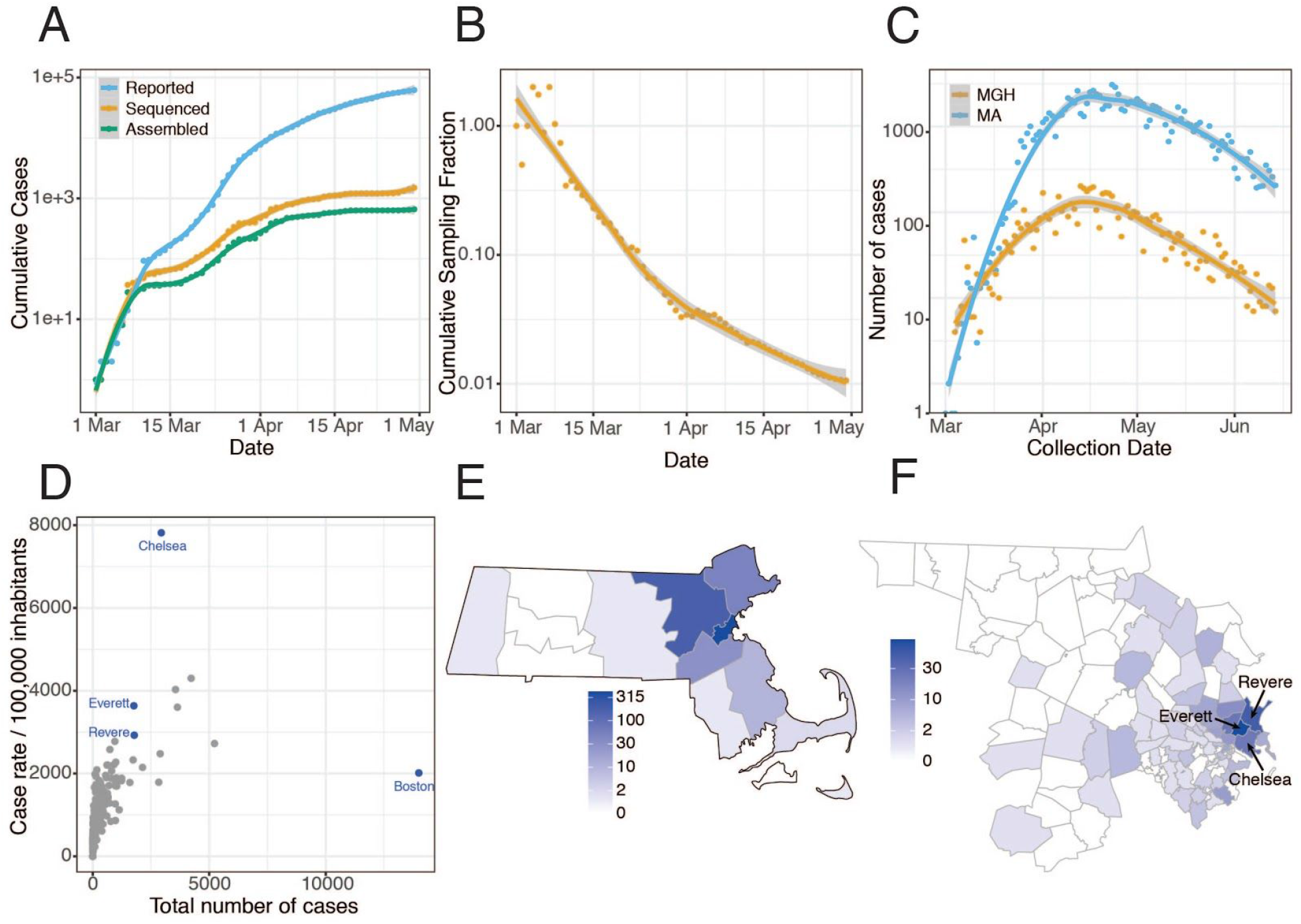
**Epidemiology of SARS-CoV-2 in Massachusetts and of sequenced viral genomes. A**. Cumulative confirmed and presumed cases reported state-wide in MA (7) from March 1 through May 1, 2020, and the number of these cases that were processed (orange) and successfully yielded complete genomes with >98% coverage (green) in this study. **B**. Cumulative proportion of all MA confirmed positive cases with complete genome sequences from unique individuals that are part of this dataset over time. **C**. Daily reported cases across MA from March 1 through June 15 statewide (blue) and at MGH (orange). **D**. Total number of cases compared to cases per **100,000** people for cities across MA. Cities in blue are highly represented in the genome dataset. **E**. Distribution of MA cases with sequenced viral genomes by county. **F**. As in E but showing only Middlesex and Suffolk counties, the two counties with the highest number of sequenced samples, by zip code. Cases associated with congregate living environments were excluded from the maps in E and F.

Viral genomes were sequenced using Illumina-based unbiased metagenomic short-read sequencing, followed by reference-guided assembly using viral-ngs 2.0.21 software (*19*) with the Wuhan-Hu-1 sequence (NC_045512.2) as the reference (Materials and Methods). We generated 778 high-quality SARS-CoV-2 assemblies (>98% complete) from 772 individuals, and an additional 72 high-quality partial genomes (>80% complete) (Fig. 1A). Genome recovery and coverage were strongly correlated with viral abundance and clinical diagnostic test results (Fig. S2 and S3). Genomes were separated from one another by a median of 6 single nucleotide polymorphisms (SNPs) (interquartile range 4-9 SNPs; range 0-85 SNPs) (Fig. S4A-B). As expected during rapid population expansion, most alleles were rare, as assessed by a strongly negative Tajima’s D statistic throughout the genome (Fig. S4C).

We constructed a phylogenetic tree from this dataset in the context of a global set of 4,011 high-quality genomes (Fig. 2A) drawn from the Global Initiative on Sharing All Influenza Data (GISAID) (Materials and Methods). Root-to-tip regression showed a clear, albeit noisy, temporal signal in our dataset, with the fitted regression model accounting for 17% of the variance in the root-to-tip distance (Fig. S5). The presence of a temporal signal means that a molecular clock can be fitted to infer the timing of ancestral branching based on SARS-CoV-2 genomes. These trees form the basis of our analysis of the Boston area epidemic.

**Fig 2.**
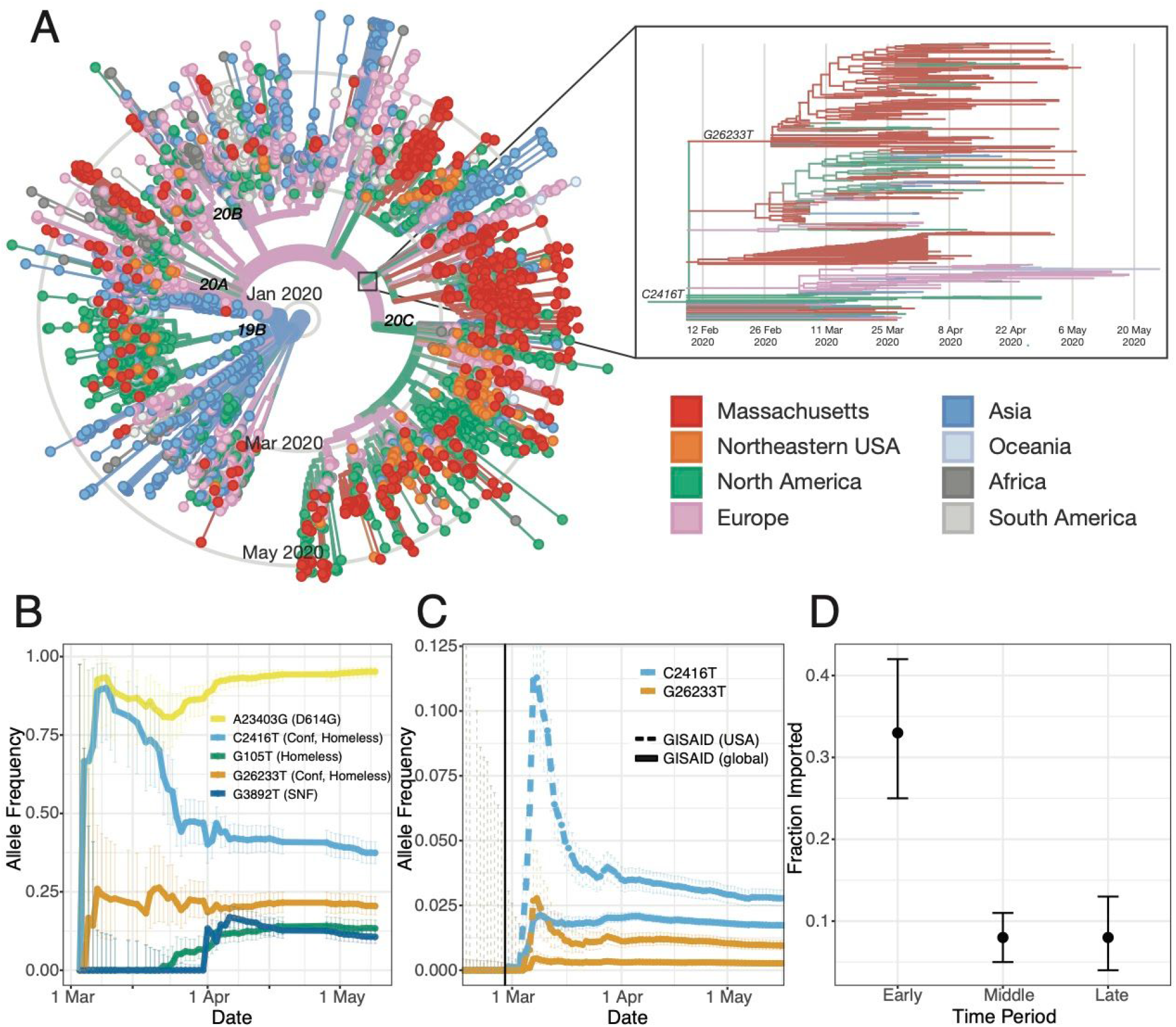
**A**. Time tree of 772 MA genomes and a global set of 4,011 high-quality genomes from GISAID. The embedded panel shows the C2416T clade in detail (outlined in gray on the main tree). To view an interactive version of this tree and for more information on specific sub-groupings within the MA dataset see auspice.broadinstitute.org. **B**. Estimated allele frequency in sequenced genomes over time for major Boston-area lineages. **C**. Frequency of the C2416T allele in 58,043 GISAID samples reported through July 14, 2020. **D**. Proportion of genomes that were inferred as imported (ancestral state as not from MA) in the early (prior to March 28, 2020), middle (March 28 - April 14, 2020) and late (after April 15, 2020) time periods of the MA epidemic.

## Introduction of SARS-CoV-2 into Massachusetts

We identified putative introductions into MA through phylogenetic analysis using an ancestral inference model (Materials and Methods). Most introductions of SARS-CoV-2 into MA occurred early in the pandemic, in March and early April, primarily from elsewhere in North America and from Europe (Table 1, Fig. 2D, and Fig. S6). We observed close connectivity between genome sequences from MA and genome sequences from elsewhere in the Northeastern USA, in particular New York (Fig. 2A). Close interstate connectivity is consistent with frequent domestic travel, which continued even after international routes were closed. The fraction of cases that were imported decreased over time (Fig. 2D), with the steepest decline during March (Fig. S6). By April 2020, the vast majority of cases resulted from local transmission rather than importation (Table 1, Fig. 2D, and Fig. S6). In total, we identified more than 80 likely introductions into MA through May 9, representing sources on four continents (Table 1).

**Table 1.** Ancestral trait inference. Results of discrete trait inference using a binary model (MA vs non-MA) and regional model (regional geographic categories) are shown, divided into date ranges representing the early, middle, and late period of the first wave of the MA epidemic.

| Region | Before March 28 | March 28 - April 15 | After April 15 |
| --- | --- | --- | --- |
| Binary model |  |  |  |
| Imported (Non-MA) | 44 | 24 | 14 |
| Not imported (MA) | 90 | 289 | 172 |
| Regional model |  |  |  |
| North America | 18 | 17 | 5 |
| Europe | 18 | 3 | 4 |
| Oceania | 1 | 0 | 0 |
| Asia | 2 | 2 | 0 |

Early diagnosed cases in MA cluster in a way consistent with their known travel and exposure history. This includes the first known COVID-19 case in MA, a traveler returning from Wuhan, China (2). Phylogenetic analysis of SARS-CoV-2 isolated from this case (named MA-1 by the CDC), based on a sample collected January 29, 2020, revealed that it clustered with others from China (Fig. S7), confirming its likely origin. Similarly, the viral sequence of the second known MA case (MA_DPH_00002), collected on March 3 from a patient who had recently traveled to Italy and Switzerland (21) clustered with European sequences (Fig. S8) and is descended from the SARS-CoV-2 genome seen in a third MA case (MA_DPH_00003), a patient who had been on the same trip. No other viral genomes in our dataset appeared to descend from these 3 cases. Thus, quarantine and contact tracing efforts appear to have prevented spread from the first known introductions into MA.

We also investigated the first cases of community transmission in MA, which occurred in Berkshire County and included several patients who had attended the same public event. Analysis of 5 viral genomes (1 complete and 4 partial) from these cases indicated that the cluster involved at least 2 introductions (Fig. S9). Four of the genomes had the same consensus sequence, indicating a common source, most likely within the United States (Fig. S10); based on the presence of the C17747) variant (i.e. a T instead of a C at position 17747), it was probably from the West Coast. We are unable to assess subsequent community transmission in Western MA since our data does not include any later Western MA samples.

## Investigation of Superspreading Events

### Spread of SARS-CoV-2 in an International Business Conference

The first large cluster of cases in MA was recognized in the context of an international business conference held in Boston from February 26 - 27 (*18*). Ultimately, more than 90 cases were diagnosed in people associated with this conference or their contacts (*22*), raising suspicion that a superspreading event had occurred there. Our dataset contains SARS-CoV-2 genomes from 28 of these cases, allowing us to look for genetic evidence of superspreading. Genetic evidence of superspreading would take the form of phylogenetic clustering of identical or highly similar viruses occurring in a narrow time window.

The signature of superspreading can indeed be seen in the conference-associated cases. All 28 genomes form a well-supported monophyletic cluster (Fig. 3A, Fig. S11) marked by the presence of the SNP C2416T (Fig. 2B-C). The parent lineage of C2416T is defined by G25563T, a lineage that was widely distributed in Europe in January and February 2020. The estimated time to the most recent common ancestor (tMRCA) for C2416T-containing genomes is February 14 (95% highest posterior density (HPD) February 4 - February 20). The C2416T variant first appears in the GISAID database in 2 French patients, ages 87 and 88, on February 29, 2020, and is absent from the 1,312 genomes in the database sampled prior to February 29 (Fig. 2C). In our dataset, all 27 C2416T-containing viruses collected prior to March 10^th^ were sampled from individuals with conference exposure, consistent with publicly available CDC genome data from MA cases from January 29 through March 7 (*24*). The rarity of C2416T in February (Fig. 2C) makes additional introductions of this allele unlikely. Taken together, this strongly suggests there was low-level community transmission of C2416T in Europe in February 2020 before the allele was introduced to Boston via a single introduction and amplified by superspreading at the conference.

**Fig 3.**
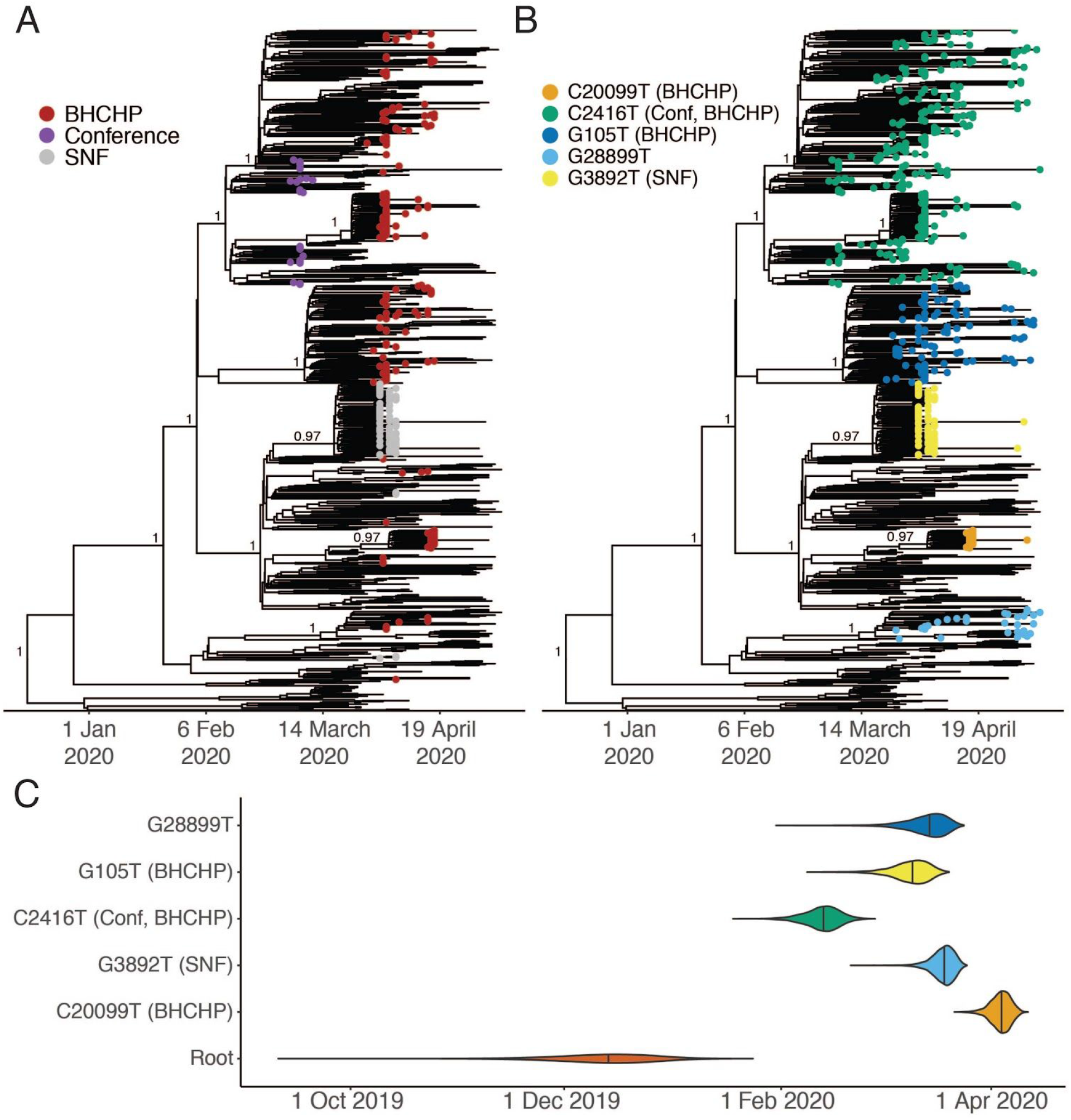
**A**. Time-measured maximum-likelihood phylogeny of 772 MA genomes. **B**. Maximum clade credibility tree with tips labeled by clade. Nodes with posterior support > 0.8 are labeled. **C**. Violin plots of tMRCA for the major Boston-area clades.

SARS-CoV-2 containing the C2416T allele subsequently spread extensively in the Boston area, representing 261/744 or 35.1% of our dataset (exclusive of known-conference associated genomes) (Fig 2B, see Sustained Local Transmission below). Beginning in early March, C2416T also appeared in multiple other US states and other countries (Fig. 2A) and increased steeply in frequency, comprising 319/11,938 (2.7%) of domestic and 937/56,118 (1.7%) of global SARS-CoV-2 genomes in GISAID collected through June 28 (Fig. 2C). The superspreading event appears to have contributed to this rise in frequency, as observed in two ways. Firstly, we identified a second variant (G26233T) that shows strong evidence of emerging during or immediately after the conference as it was first seen in 7 of 28 individuals with known exposure to the conference, including in one sample from a conference attendee at intermediate frequency (26%). C2416T/G26233T was subsequently exported from Boston to several US states, including Virginia, North Carolina, and Texas, and to other countries, including Australia, Sweden, and Slovakia (Fig. 2A, Fig. S12A-B), with evidence of community spread in Virginia, Australia, and Michigan. Secondly, we assessed the extent to which US spread of C2416T could be due to additional importations from Europe. Two European sub-lineages (C2416T/G8371T and C2416T/G20578T) are extremely rare in the United States: 0/73 genomes and 1/228 genomes, respectively, compared with 24/197 genomes containing the C2416T/G26233T mutations (Fig. S12C-D). This, along with epidemiological data connecting multiple conference-linked cases to other US states (*25*-*28*), suggests that most C2416T viruses in the US likely derive from this initial introduction. However, we cannot estimate the absolute number of individuals involved as sequenced genomes are not a random sample of cases and US state-level data is highly incomplete at this time.

### Spread of SARS-CoV-2 In Homeless Shelter Guests and Staff

To support public health investigations of transmission in high priority populations, we analyzed the introduction and spread of SARS-CoV-2 in homeless shelter guests and staff served by the Boston Health Care for the Homeless Program (BHCHP). Samples were collected in March and April 2020 and included those collected during universal screening at Boston’s largest homeless shelter(*16*). From these samples, we assembled and inferred a phylogeny from 193 SARS-CoV-2 genomes (Fig. 2A, Fig. 3A and Fig. 3B). We identified at least 7 introductions into the BHCHP population, including 4 that resulted in clusters containing 20 or more highly similar viral genomes (Fig. 4A and Fig. 4C); a phylogenetic signature consistent with superspreading. Two of the clusters were of genomes descended from the C2416T lineage: of the 193 genomes, 105 (54.4%) contained C2416T, and 54 of these 105 (51.4%) additionally contained G26233T, demonstrating that BHCHP guests and staff were affected by community transmission resulting from conference-associated amplification and spread of SARS-CoV-2.

**Fig 4.**
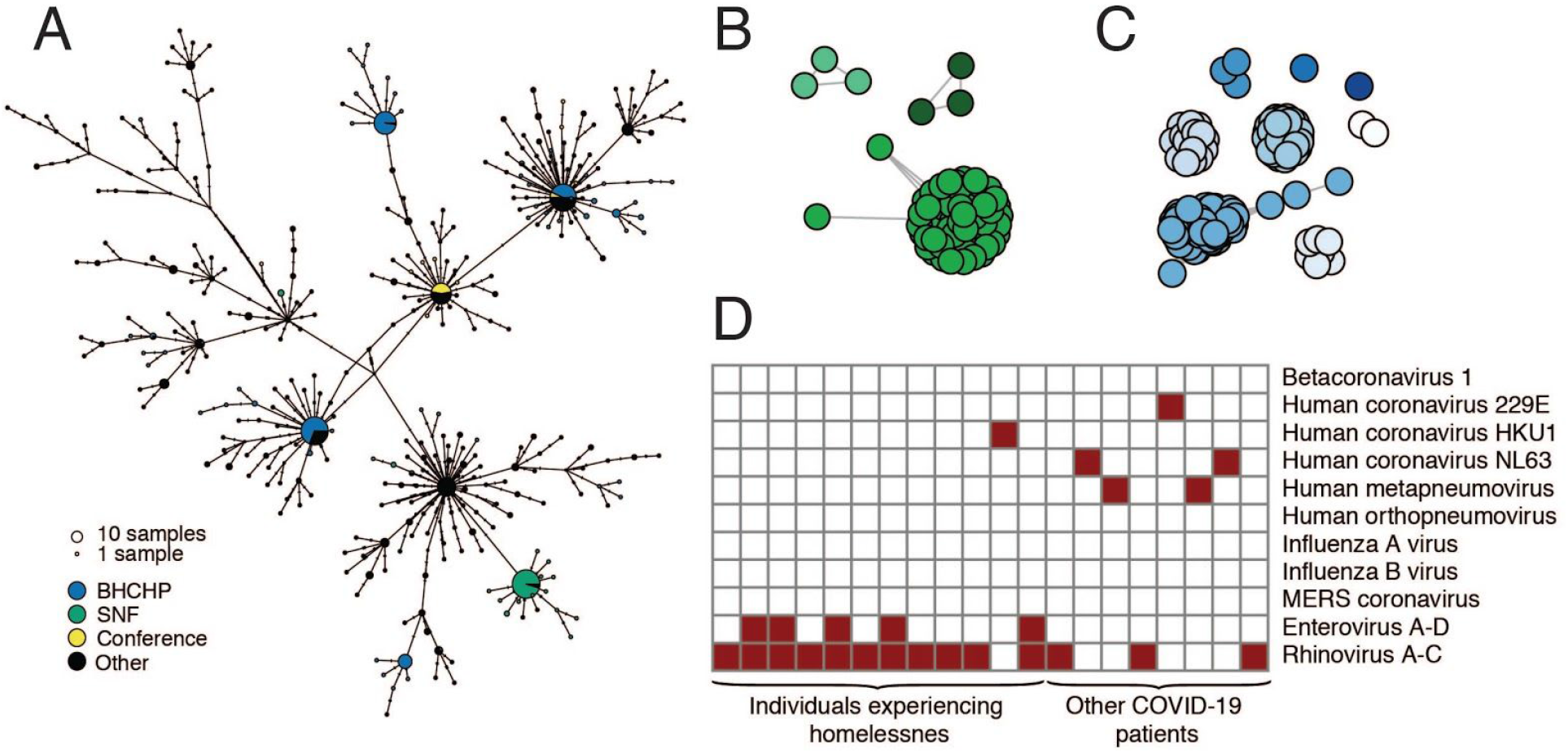
**SARS-CoV-2 superspreading events. A**. Haplotype network of SARS-CoV-2 haplotypes in the MA dataset with major known superspreading events highlighted. **B, C**. Gene graphs showing clusters of highly similar sequences among viral genomes from the SNF **(B)** and BHCHP **(C)** cohorts. Sequences are clustered when they are separated by < 4 SNPs, and the lengths of lines between points reflect genetic distance. **D**. Detection of common respiratory viruses from metagenomic sequencing data. Samples with >10 reads mapped to at least 1 of these viruses using Kraken2 are shown in red. Enterovirus and Rhinovirus species have been grouped due to difficulty in discriminating at the sequence level.

### Spread of SARS-CoV-2 In a Skilled Nursing Facility

We also investigated cases in another vulnerable population, cases that were involved in a superspreading event at a skilled nursing facility (SNF) in the Boston area. Prior to a planned relocation of residents in early April, universal screening detected SARS-CoV-2 in 82/97 (85%) of the residents and 36/97 (37%) of the staff by RT-qPCR (*29*). We assembled 83 SARS-CoV-2 genomes from these individuals, 75 of which comprised a single cluster of closely related genomes (59 identical), all containing a G3892T mutation (posterior support of 1 in maximum clade credibility tree, Fig. 3A and Fig. 3B). The paucity of genetic variation within the cluster implies that introduction into the facility had been recent and from a single predominant source (Fig. 4A). Consistent with this, the median tMRCA for sequences in the cluster was March 20 (Fig. 3C, 95 % HPD: March 13 - March 24, 2020). The estimated tMRCA along with the high proportion (30/45) of residents who tested negative on April 1, 2020, but were found to be positive 5 days later (*29*) suggests rapid spread within the facility in late March and early April 2020.

The genetic diversity in the SNF cluster is strikingly low even under the assumption of recent transmission from a single source. The 18 mutations seen in the cluster is significantly lower than what we would expect based on the conference cluster (p = 0.019), which occurred over a similarly short time window, and much lower than the 30 mutations expected under a simple model of SARS-CoV-2 substitution (p = 0.011, Materials and Methods). The low genetic diversity in this cluster might simply result from low diversity in the index patient, but could also reflect differences in the transmission process at work. For example, if more virions than usual were transmitted to each SNF case, then the resulting infections would more often have the same consensus genome as the index case.

In addition to the major SNF cluster, two other introductions, each containing three genomes, can also be seen among the patients and staff in the SNF (Fig. 3A and Fig. 4B). There is strong phylogenetic support for each of these two separate introductions (Fig 3A). The observation that one introduction led to massive spread, while the other two did not, is consistent with reported overdispersion of secondary infections in several coronaviruses (*8*, *12*, *30*, *31*); that is, there is more variance in the number of secondary infections caused by each case than expected from a random Poisson process. Although one clade predominated within this SNF, the occurrence of at least three independent introductions underscores the high risk of introduction into a single facility (*32*). These introductions occurred despite strict infection control policies—including a restriction on visitors (*33*), universal masking for all staff, masking for all residents when leaving their rooms, and vigilance with hand hygiene—in place for at least two weeks before the first detected infection (*29*).

### Investigation of potential nosocomial outbreaks

We investigated two case clusters at MGH for which the Infection Control Unit raised suspicions of a nosocomial outbreak. In the first cluster, two patients in the same hospital ward tested positive for SARS-CoV-2 during their hospital stay after testing negative at the time of admission. In the second cluster, five patients who received care in a speciality ward were diagnosed with SARS-CoV-2 infections over a period of several days. For each cluster, complete genomes (2 of 2 from the first cluster and 4 of 5 from the second cluster) were inconsistent with a common ancestor during the period of hospitalization (Fig. S13). We therefore rejected the hypothesis that the individuals in each cluster were part of the same transmission chain, although we cannot exclude the possibility of nosocomial transmission per se because independent introductions from multiple asymptomatic staff could theoretically have occurred.

## Sustained Local Transmission

Our dataset covers March 3 through May 9, an interval that spans the beginning, peak, and initial decline of the first wave of the epidemic in MA (Fig. 1C). Several clades established early in the Boston-area outbreak showed continued community transmission throughout that period (Table 2, Fig. 3A-B), with the lineage containing C2416T, associated with the conference, being the largest. The C2416T lineage was likely the first of these clades imported into Boston (median estimated tMRCA, February 14, 2020; 95% HPD February 4 - 20, 2020) (Fig. 3C). The other four major lineages (G3892T, G105T, G28899T, and C20099T) appeared to enter the region between March and early April 2020. Consistent with a larger global trend (*34*, *35*), we observed a rise in frequency of viruses harboring the D614G amino acid polymorphism, conferred by a SNP at nucleotide 23,403 in the Wuhan reference strain, which rose to near-fixation in MA by the end of the study period (Fig. 2B) and is present in all of the dominant lineages.

**Table 2:** Major Boston-area lineages identified by lineage-defining mutation.

| Lineage | Root | C20099T | G3892T | C2416T | G105T | G28899T |
| --- | --- | --- | --- | --- | --- | --- |
| Number of Genomes | 772 | 21 | 77 | 288 | 98 | 34 |
| Epidemiology |  | BHCHP | SNF | Conference, BHCHP | BHCHP |  |
| Amino Acid substitution |  | ORF1b: A2211V;<br>NSP15: A160V | ORF1a: E1209D;<br>NSP3: E391D |  |  | N: R56I, ORF14: E56* |
| Median tMRCA (95% HPD) | 2019-12-15<br>(2019-11-20 - 2020-01-04) | 2020-04-04<br>(2020-03-30 - 2020-04-08) | 2020-03-19<br>(2020-03-13 - 2020-03-23) | 2020-02-14<br>(2020-02-04 - 2020-02-20) | 2020-03-10<br>(2020-03-01 - 2020-03-16) | 2020-03-15<br>(2020-03-04 - 2020-03-21) |

Based on tMRCA estimates for major Boston-area clades, we do not find evidence of undetected, or “cryptic,” transmission before mid-February, although small outbreaks may have gone undetected. In addition to the lineages reported here, none of the importation events we inferred (Table 1) occurred prior to known cases, although testing for SARS-CoV-2 in MA was restricted to a narrow definition prior to established community spread (*36*). In particular, additional isolated events similar to the MA-1 importation may have occurred and escaped detection with the current resolution of sampling.

Phylogenetic data, when labeled by patient zip code (Fig. S14), reveal that all major lineages, including conference-associated viruses, were circulating in the Boston-area communities of Chelsea, Revere, and Everett, which were among the most heavily affected communities in the state (Fig. 1D). Thus, while viral lineages entered and were amplified by distinct mechanisms, cases rapidly spread between communities (Fig. S15). C2416T was the most common lineage in the Boston area throughout the study period and across sampling sites (Fig. S16). By the end of that period, the cumulative allele frequency of C2416T in our dataset, exclusive of conference- and SNF-associated samples, was 46.4% (194/418) in Suffolk, 30.1% (31/103) in Middlesex, 30.0% in Essex (12/40), and 40.9% (9/22) in Norfolk counties. The conference superspreading event likely had an outsized effect because it occurred early in the pandemic (Fig. S17). Extensive spread within the Boston area likely then contributed to the rise in frequency of C2416T and C26233T in the United States and worldwide (Fig. 2B-C).

### Respiratory Viral Coinfections

The metagenomic approach we used for sequencing SARS-CoV-2 enabled us to screen for respiratory viral co-infections in patients with COVID-19. We found other respiratory viruses in 20/1431 (1.4%) of COVID-19 cases (Fig. 4D) and confirmed these results using the BioFire FilmArray Respiratory Panel (Fig. S18). The most common co-infecting viruses were Rhinovirus/Enterovirus. The rarity of co-infections (lower than reported in an early dataset from California (*37*)) likely reflects the timing of the SARS-CoV-2 epidemic in MA, which began near the end of the influenza season; weekly data from MADPH show rapidly declining influenza activity in March and April 2020 (*38*). We observed a higher rate of co-infection with other respiratory pathogens among BHCHP clients and staff (12/314) than in the other samples in our dataset (8/1117) (p = 0.0002, Fisher’s exact test), consistent with an increased epidemiological risk in this population.

## Conclusions

We present here an analysis of SARS-CoV-2 genomic epidemiology primarily in the Boston area, which was severely affected early in the US COVID-19 epidemic. Through dense sampling of the early phase of the epidemic we show the frequency of importation events—over 80 independent introductions—and the impact of early superspreading events in driving amplification and community transmission, likely accelerating the transition from containment to mitigation strategies.

Besides better understanding of outbreak dynamics, viral sequencing and phylogenetic analysis can also provide immediately actionable insights. In the current study, we were able to rule out linked nosocomial spread in two episodes, reassuring hospital management that a failure of infection control practice in these wards had not led to a nosocomial cluster, and showed that despite multiple introductions of SARS-CoV-2 into a SNF, one introduction was responsible for 90% of cases. Real-time genomic epidemiology may be increasingly valuable as schools and workplaces navigate the challenges of reopening, as it can help distinguish between local outbreaks within institutions and introductions from outside.

The relatively narrow surveillance definition for SARS-CoV-2 in MA until March 4 may have limited identification of other early introductions or delayed detection of some individuals who did not meet testing criteria. Similarly, the dataset is not a random sample over time, comprised instead largely of cases that fell in the MGH catchment area or that were sampled from particular subpopulations to gain insight into local epidemiology.

Our findings repeatedly highlight the close relationships between seemingly disconnected groups and populations: viruses from international business travel seeded major outbreaks among individuals experiencing homelessness, spread throughout the Boston area, and were exported to other domestic and international sites. It also illustrates the role of chance in the trajectory of an epidemic: a single introduction had an outsize effect on subsequent transmission because it was unfortunately amplified by superspreading in a highly mobile population very early in the outbreak, before many precautions were put in place and when its effects would be further amplified by exponential growth. By contrast, other early introductions led to very little onward transmission, and another superspreading event in a SNF, while devastating to the residents, had little large-scale effect because it occurred later and in a more isolated population. This study provides direct evidence that superspreading events may profoundly alter the course of an epidemic and implies that prevention, detection, and mitigation of such events should be a priority for public health efforts.

## Data Availability

Interactive phylogenetic trees are available at auspice.broadinstitute.org. Assembled genomes and raw metagenomic reads from this dataset have been deposited at NCBI Genbank and SRA databases under BioProject PRJNA622837 with NIAID Data Sharing policy. Experimental protocols are publicly available on Benchling and can be accessed at the link below.

https://benchling.com/sabetilab/f_/gaLGu5X9-sabeti_group_sars-cov-2_metagenomic_sequencing_protocols/

https://auspice.broadinstitute.org

## Acknowledgements

We gratefully acknowledge the microbiology lab staff at MGH and DPH and all members of the COVID-19 emergency response efforts at MGH, BHCHP, and MADPH. We also thank Hayden C. Metsky for valuable feedback.

## Author contributions

### Data and materials availability

Sequences and genome assembly data are publicly available in the Broad Institute’s <u>Terra</u> platform in a <u>featured workspace for COVID-19</u>. Researchers can use this workspace to reproduce analyses described here or perform similar analyses on their own viral sequence data. Assembled genomes and raw metagenomic reads from this dataset have been deposited at NCBI’s Genbank and SRA databases under BioProject PRJNA622837 in accordance with <u>NIAID’s Data Sharing policy</u> and will soon be available to visualize on nextstrain.ora/ncov. Experimental protocols are publicly available on Benchling and can be accessed here: https://benchling.com/sabetilab/f_/aaLGu5X9-sabeti_group_sars-cov-2_metaaenomic_sequencing_protocols/.

## List of Supplementary Materials

Materials and Methods

Table S1 – S3

Fig S1 – S18

References (1 – 54)

## Supplementary Materials

### MATERIALS AND METHODS

#### Sample collections

This study was approved by the Partners Institutional Review Board under protocol 2019P003305 and MDPH IRB 00000701. We obtained samples and selected metadata from the MGH Microbiology Laboratory and MADPH under a waiver of consent for viral genomic sequencing. Samples were tested for SARS-CoV-2 by RT-qPCR. Samples that tested positive were eligible to be included.

Archived samples obtained from the MGH Microbiology Laboratory included nasopharyngeal (NP) swabs from five sources 1) all available cases prior to March 8, 2020, 2) all available samples from a skilled nursing facility in the Boston area (29), 3) samples from April 1 through April 14 from the MGH Respiratory Illness Clinic (RIC), established in Chelsea, MA, 4) samples from MGH Infection Control Unit investigations, and 5) samples drawn from the general pool of available samples tested by the MGH Microbiology Laboratory during the period from March 4 through May 9, 2020. Archived samples obtained from MADPH included NP swabs from 1) all available samples representing the first two known travel-associated introductions and the Berkshire County cluster and 2) all available samples submitted to MADPH from Boston Healthcare for the Homeless Program (BHCHP) from Mar 19 through April 18, a period that included universal screening (*16*).

#### Annotation of Cases

Epidemiological data on exposure and geography were obtained from medical record review (MGH) or collected by the DPH laboratory in the process of clinical testing. Zip code and county-level data were available for most samples from MGH. County-level data was available from DPH samples. Individuals who participated in the conference or who had known direct contact with attendees of the conference were deemed conference-associated (n = 28). One additional patient reported staying at the conference hotel but was diagnosed with COVID-19 over 1 month later;their exposure was considered unlikely to be due to the conference.

#### Viral sequencing

Samples were received at the Broad Institute as viral transport medium, universal transport medium, or molecular transport medium from NP swabs. In accordance with institutional biosafety committee approvals, samples were inactivated with Buffer AVL (Qiagen) or other chaotropic salt solution prior to extraction. RNA was extracted from 200uL of transport medium using either the QiAmp Viral RNA Mini Kit (Qiagen), or the MagMAX mirVana Total RNA Isolation kit on a KingFisher Flex automated extraction instrument (Thermo Fisher Scientific). Residual DNA was removed from the extracted material using TURBO DNase (Thermo Fisher Scientific).

Human ribosomal RNA was depleted using a ssDNA probe-based RNase H depletion method as previously described (*39, 40*), or with the Ribo-Zero Plus rRNA Depletion Kit (Illumina). Unique ERCC RNA spike-ins were added to each sample as a quality control measure to track and mitigate potential cross contamination or downstream sample preparation issues. First and second strand cDNA was synthesized using either superscript III or IV Reverse Transcriptase (Thermo Fisher Scientific), and sequencing libraries were prepared with the Nextera XT or TruSeq RNA Library Prep kits as previously described (*39, 40*). Libraries were sequenced using Illumina MiSeq, HiSeq, NextSeq, or NovaSeq machines with 100-nucleotide paired-end reads. samples were extracted, prepared, and sequenced at the Broad Institute, Cambridge, MA, USA. The rRNA depletion, cDNA synthesis, and library construction protocols used in this study are publicly available on Benchling and can be found here: https://benchling.com/sabetilab/f_/aaLGu5X9-sabeti_group_sars-cov-2_metaaenomic_sequencing_protocols/.

#### Genomic data analysis

We conducted all analyses using viral-ngs 2.0.21 on the Terra platform (app.terra.bioV All of the workflows named below are publicly available via the Dockstore Tool Repository Service (dockstore.oro/oroanizations/Broadlnstitute/collections/pgs). We demultiplexed individual libraries using the *demux_only* workflow for each lane of each flowcell, removed reads mapping to the human genome and to other known technical contaminants (e.g. sequencing adapters) using *deplete_only* (with bwaDbs=[“gs://pathogen-public-dbs/v0/hg19.bwa_idx.tar.zst”] and blastDbs=[“gs://pathogen-public-dbs/v0/GRCh37.68_ncRNA.fasta.zst”, “gs://pathogen-public-dbs/vO/hybsel_probe_adapters.fasta”]), and performed reference-based assembly using *assemble_refbased* (once per sample, with all sequencing replicates merged in the read_unmapped_bams input and with a reference_fasta taken from https://www.ncbi.nlm.nih.gov/nuccore/NC_045512.2?report=fasta) We ran *assemble_refbased* on 1970 read set inputs spanning 1535 distinct samples (inclusive of controls).

We used the following stringent criteria to excluded any sample where i) fewer than 50,000 cleaned reads were obtained; ii) the proportion of reads mapping to the internal control (IC) sequence (ERCC spike-in) was >3 standard deviations from the mean observed for that IC sequence across all sequencing batches; iii) replicate genomes—where available—had 2 or more discordant SNPs or 1 or more discordant indels; iv) the number of normalized reads mapping to the SARS-CoV-2 genome was less than that observed in the highest negative control from the same sequencing batch. From the 1196 patient samples after filtering we obtained 850 assemblies with unambiguous consensus calls over at least 80% of the SARS-CoV-2 genome, and 778 with over unambiguous consensus calls over at least 98% of the SARS-CoV-2 genome, of which 772 were from unique individuals. We submitted 633 read sets to NCBI SRA and 837 genomes with at least 80% completeness to NCBI Genbank (using the *genbank* workflow). We used the 772 high-quality assemblies from unique individuals for the phylogenetic analyses described.

Failure to produce a SARS-CoV-2 genome from a PCR-positive sample may have been due to low viral titer, RNA degradation due to lack of sufficient cold chain, or technical sample handling issues (e.g. improper swab technique). Samples which failed to produce a genome at the first attempt were not further investigated at this time. To confirm the quality of our assemblies and mitigate any potential contamination we performed replicate library preparation and sequencing from RNA for 10% of samples. Among those samples that assembled a complete genome in both replicates, consensus-level genomes were identical.

Allele frequency was estimated as the proportion of derived / (derived + ancestral) versions of the allele. A 95% confidence interval was estimated for the proportion using the binomial distribution. The frequency of the iSNV for MA_MGH_00427 was calculated from 2 libraries; 50 reads contained the derived T allele and 146 reads contained the ancestral G allele based on the aligned reads from the viral-ngs pipeline (as described above).

#### Phylogenetic tree reconstruction

We constructed phylogenetic maximum likelihood (ML) and time trees with associated visualizations using the Augur pipeline (*augur_with_assemblies*). We used SARS-CoV-2-specific procedures taken from aithub.com/nextstrain/ncov. specifically setting the clock rate to 0.0008 +/- 0.0004, rooting the tree using the reference genome, and using the nextstrain site-masking and clade-definition files. In addition to our 772 genomes from unique individuals from Massachusetts, we included a global comparison set of 4,011 genomes subsampled from a download from the GISAID database on 15 June, 2020. These 4,011 genomes contain at most 50 representatives from each state or province in North America plus at most 50 representatives from each country outside of North America. Random subsampling was biased towards genomes genetically close to our focal set of genomes, using the distance matrix calculator at github.com/nextstrain/ncov/blob/master/scripts/priorities.py. The resulting augur output is visualizable on auspice.us or can be incorporated in custom deployments using Google Cloud Run using our template (github.com/dpark01/auspice-private-template); this template is used to showcase our data at auspice.broadinstitute.org.

We also conducted additional analysis of the genomes sequenced in this study. We aligned the set of 772 genomes using MAFFT v7.471(47) and trimmed 5’ and 3’ (first 265 and last 228 bases) UTRs from the alignment in R (*42*). To estimate the root-to-tip distance, we constructed ML phylogenetic trees using PhyML(*43*) v3.3.20190909 with default parameters using the MAFFT alignment of 772 genomes. We used TempEst (*44*) v.1.5.3 and selected the best-fitting root as identified using a heuristic residual mean squared function. To estimate branch support in maximum-likelihood phylogenies, we used IQ-Tree (*45*) with the ultrafast bootstrap and 10,000 bootstrap samples.

To construct Bayesian time-trees, we used BEAST 2.6.2 with a general time reversible substitution model with 4 rate categories drawn from a gamma distribution (GTR4G), a strict clock, coalescent exponential tree prior, a uniform [-inf, inf] prior for the clock rate, a 1/x [-inf, inf] prior for the coalescent effective population size; and a laplace [-inf, inf] prior for the growth rate. We ran the MCMC chain in BEAST2 for 100 million steps and thinned the chain by recording samples every 1000 steps. The first 30% of samples were discarded prior to calculating summary statistics from the posterior. We used TreeAnnotator v2.6.2 to construct maximum clade credibility trees with a burn-in percentage of 30%. We also compared a Hasegawa-Yoshino-Gawa substitution model with kappa = 2 and with 4 rate categories drawn from the gamma distribution (HKY4G) and ran this chain for 100 million steps using the same thinning and burn-in described for the GTR4G model.

#### Detection of respiratory virus co-infection

We used Kraken2 (*46*) to identify other viral taxa present in NP swab samples from COVID positive patients, excluding those removed by filters i and ii described above. To do so, we ran the *classify_single* workflow on all reads from all samples (with kraken2_db_tgz=”gs://pathogen-public-dbs/v1/kraken2-broad-20200505.tar.zst”, krona_taxonomy_db_kraken2_tgz=”gs://pathogen-public-dbs/v1/krona. taxonomy-20200505, tab. zst”, ncbi_taxdump_tgz=”gs://pathogen-public-dbs/v1/taxdump-20200505.tar.gz”, trim_clip_db=”gs://pathogen-public-dbs/v0/contaminants.clip_db.fasta”, spikein_db=”gs://pathogen-public-dbs/v0/ERCC_96_nopolyA.fasta”). Our kraken2 database was constructed on 5 May, 2020, with the *kraken2_build* workflow (with standard_libraries=[“archaea”, “bacteria”, “plasmid”, “viral”, “human”, “fungi”, “protozoa”, “UniVec_Core”] and custom_libraries=[“gs://pathogen-public-dbs/v1/Hybsel_Viruses-20170523.2.fa.zst”, “gs://pathogen-public-dbs/v1/ercc_spike-ins-20170523.fa”]). The resulting per-sample outputs were run through the *merge_metagenomics* workflow and the resulting hits were filtered down to 20 common respiratory viruses of interest (adenovirus, HCoV-229E, HCoV-HKU1, HCoV-NL63, betacoronavirus 1, parainfluenza 1, parainfluenza 2, parainfluenza 3, Parainfluenza 4, enterovirus A, enterovirus B, enterovirus C, enterovirus D, influenza A, influenza B, human metapneumovirus, respiratory syncytial virus, SARS-CoV, MERS-CoV, human rhinovirus) using a threshold of 10 reads to identify a putative co-infection. We independently confirmed the presence of viral co-infections identified in the metagenomic sequencing data using the BioFire FilmAssay Respiratory Panel, performed at the MADPH or MGH Microbiology Laboratory. Three samples from early in the pandemic, for which no additional sample remained, were not tested.

#### Identifying viral importation events

For ancestral state inference, we inferred a state of “MA” vs “non-MA” using the augur pipeline(47). Cases whose ancestral state was inferred as non-MA with high confidence (>0.95) were considered imported cases. Conference-associated and nursing facility samples were excluded from the importation analysis.

#### Haplotype Network Reconstruction

Haplotype networks were visualized using the software tool PopART v1.7 (*48*). The assembled sequences were aligned against NC_045512.2 and the first 268bp at the 5’ end and 230bp at the 3’ end (UTR regions) were removed from the alignment. A nexus-format input file for PopART was created using a Python script to consolidate sequence information with metadata classifications. This script is available at http://www.github.com/broadinstitute/trepository], A TCS network of the sequences (*49*) was constructed in PopART. Regions where any sequence had ambiguous bases were masked. For the construction of haplotype networks in Figure 4, one sample, MA_MGH_00090, was removed to prevent masking of the G3892T variant. For the displayed haplotype networks, the area of the circle corresponds to how many verbatim-identical sequences (after masking) bin together as the same haplotype. The hash marks on the edges indicate the SNP distance between sequence haplotypes (1 mark=1 SNP apart). Gene graphs were constructed using pairwise distance matrices computed on aligned SARS-CoV-2 genomes and clustered using the R package adegenet (*50*).

#### SNF genetic diversity analysis

For this analysis, the main SNF cluster was restricted to samples collected before April 15, 2020, and the conference cluster to samples collected before March 8, 2020. We assumed that the number of transmissions was the minimum possible (one fewer than the number of samples in the cluster). The p-value for the comparison between the clusters assessed the probability that the observed numbers of mutations were produced by Poisson processes with the same value of λ, using the R function poisson.test (in the stats package v3.6.2). For the expected number of mutations, we assumed that substitutions occur predominantly during the transmission bottleneck and calculated the expected rate based on a generation time of 5.0 days {51) and a mutation rate of 1.0 × 10^-3^/bp/year (Fig. S5C), which together yield an expectation of 0.41 substitutions/transmission.

#### Epidemiological and demographic data analysis

We downloaded publicly available daily and weekly data on cases of SARS-CoV-2 in MA for the period January 1 - August 1 from the website of the MADPH (https://www.mass.aov/info-details/covid-19-response-reporting). This data included cases by day, cases by county over time, and cases involving congregate living facilities and staff. We compiled detailed case statistics by exposure category using the press releases reporting early case totals and exposure available on the MADPH website. During the study period, an additional case from February 6, 2020, was added to MADPH tallies. This case was missing detailed case information such as exposure category and was not included in early press releases from MADPH; it was therefore excluded from the tallies of cases by exposure category and estimates of the sampling proportion, but included in total case counts over time as reported in the main text to incorporate the most recent tallies. To calculate the cumulative proportion of alleles by county, conference-associated and SNF-associated individuals were removed and the cumulative allele frequency through the end of the study period was calculated for each of the four counties with the largest numbers of genomes (Suffolk, Middlesex, Essex, and Norfolk). To calculate the proportion of domestic and global sequences from the GISAID database, a multiple sequence alignment of 58,043 complete GISAID genomes was downloaded on July 14 2020 and the percentage of ancestral and derived alleles was extracted from the alignment and plotted by geographic category.

We used R (42), Bioconductor (52), ggplot2, tidyverse (53), and ggtree (54) to clean and plot data and trees, and choroplethr to draw maps.

**Fig S1.**
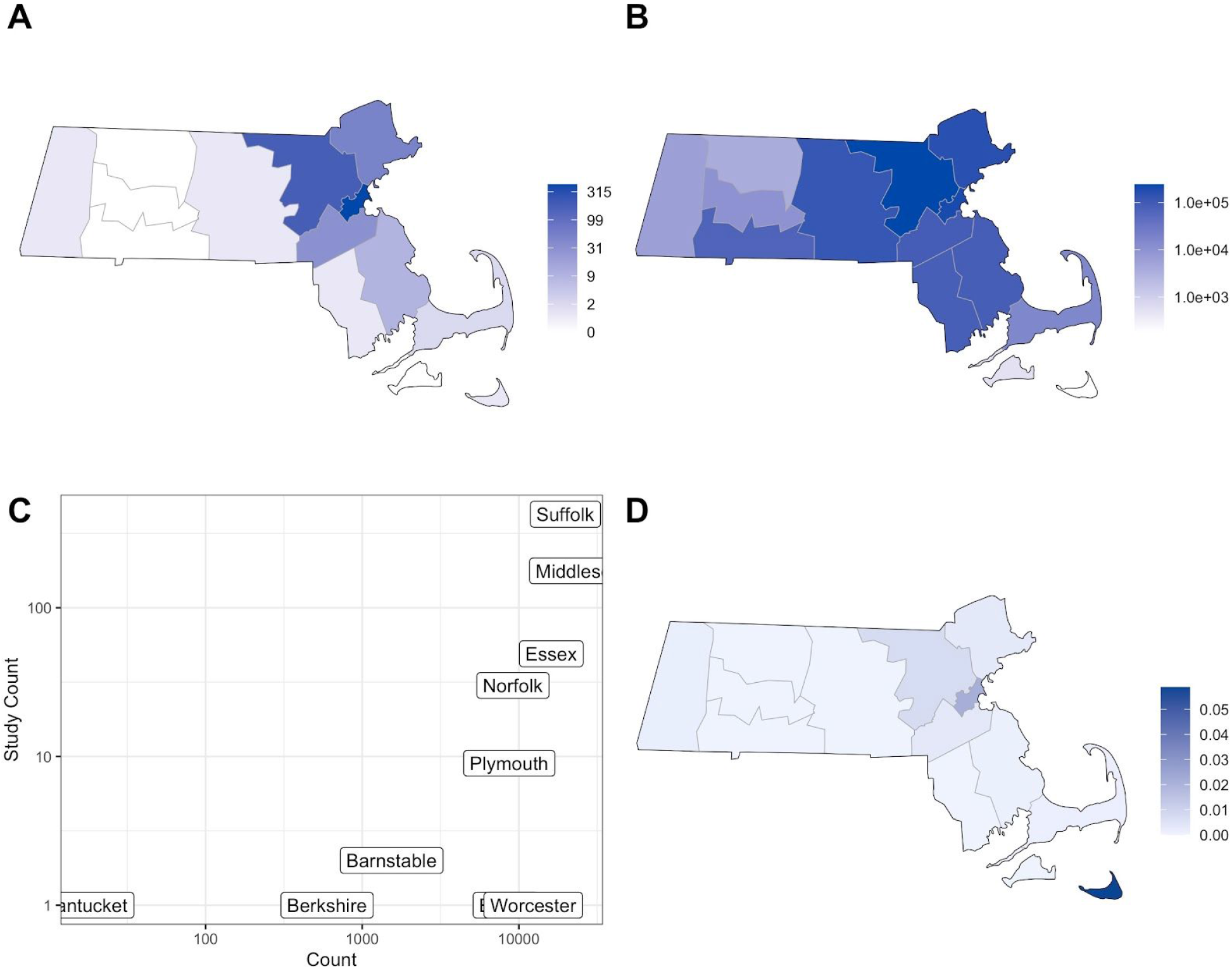
**A**. Counts of complete genomes reported in this study, by county. **B**. Case counts by county reported by MADPH through July 1, 2020. **C**. Scatterplot of counts of complete genomes in this study vs. MADPH-reported cases through July 1, 2020. **D**. Sampling proportion by county (fraction of complete genomes sequenced in this study out of total cases by county reported to MADPH through July 1, 2020).

**Fig S2.**
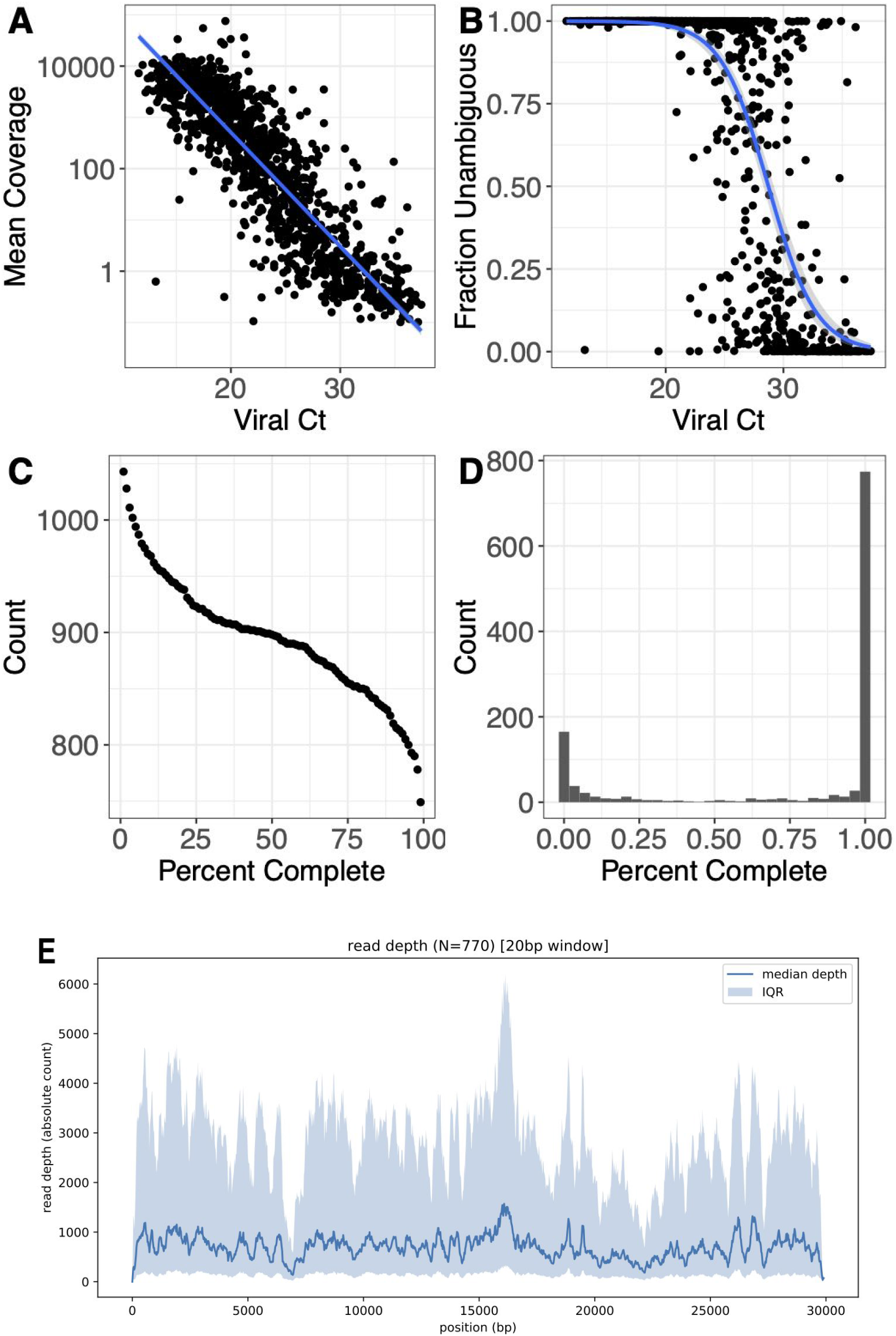
**A**. Mean coverage (on a log scale) vs. viral C_t_ for all samples included in the study. A linear regression fit is shown in blue. **B**. Fraction of the genome that is complete is shown vs. viral C_t_. A C_t_ < 28 was strongly associated with recovery of a complete virus genome. Fit from a logistic regression model is shown in blue. **C**. The numbers of genomes at given thresholds of completeness are displayed. **D**. Histogram of the numbers of genomes at different thresholds of completeness. **E**. Combined coverage across sequenced SARS-CoV-2 genomes, [next page]

**Fig S3.**
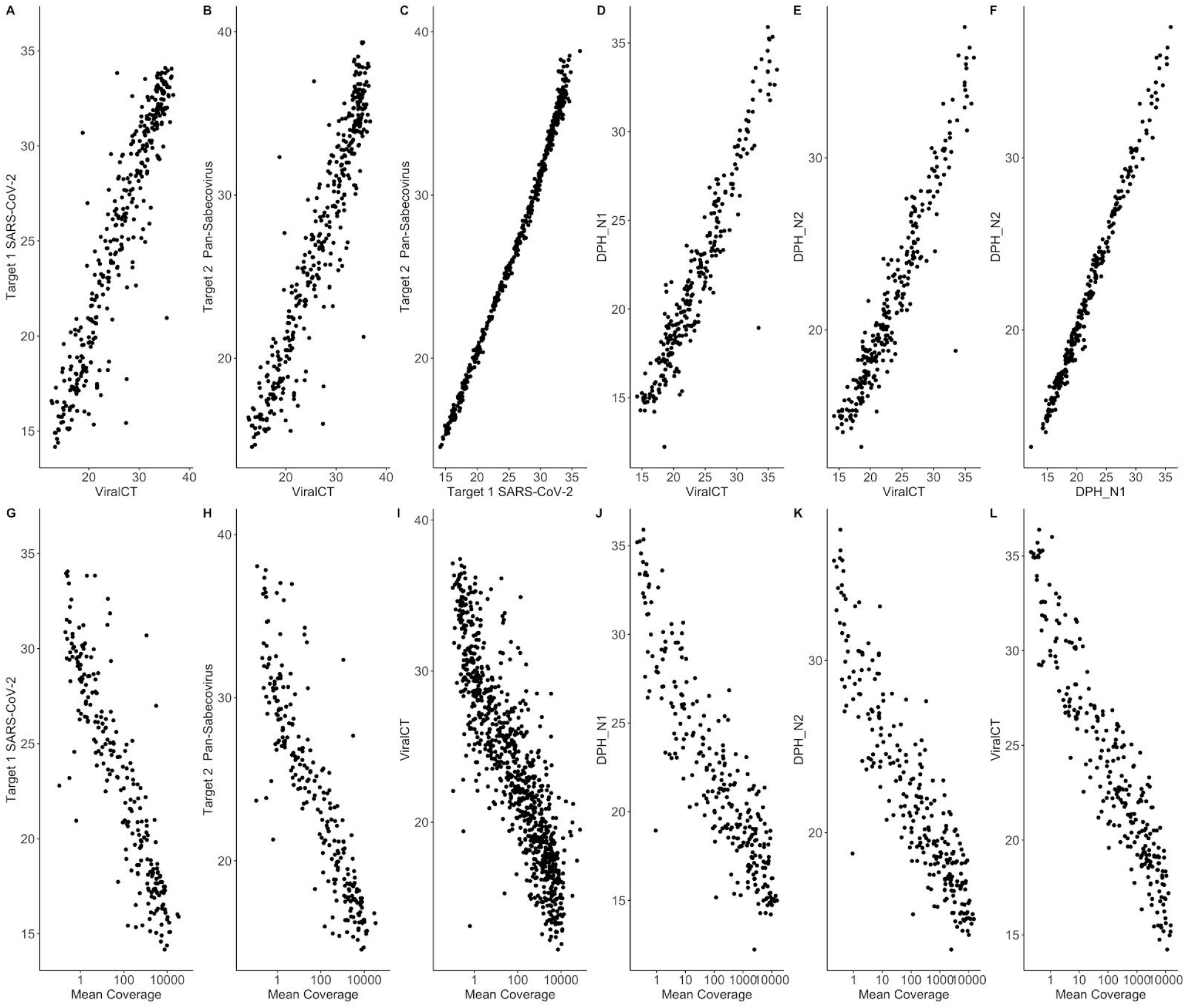
**A**. Scatterplot of MGH Roche cobas 6800 instrument PCR Ct values for SARS-CoV-2 target vs. quantification prior to library construction. **B**. Scatterplot of MGH Roche cobas 6800 instrument PCR C_t_ values for Pan SARS target vs. quantification prior to library construction. **C**. Scatterplot of Roche cobas 6800 PCR C_t_ targets. **D**. Scatterplot of DPH N1 assay vs. quantification prior to library construction. **E**. Scatterplot of DPH N2 assay vs. quantification prior to library construction. **F**. Scatterplot of DPH N1 vs. N2 targets. **G**. Scatterplot of MGH Roche cobas 6800 instrument PCR C_t_ values for SARS-CoV-2 target vs. mean coverage (log 10 scale). **H**. Scatterplot of MGH Roche cobas 6800 instrument PCR C_t_ values for Pan SARS target vs. mean coverage (log 10 scale). I. Quantification prior to sequencing vs. mean coverage (log 10 scale) for MGH samples. **J**. Scatterplot of DPH N1 assay vs. mean coverage (log 10 scale). **K**. Scatterplot of DPH N2 assay vs. mean coverage (log 10 scale). **L**. Quantification prior to sequencing vs. mean coverage (log 10 scale) for DPH samples.

**Fig S4.**
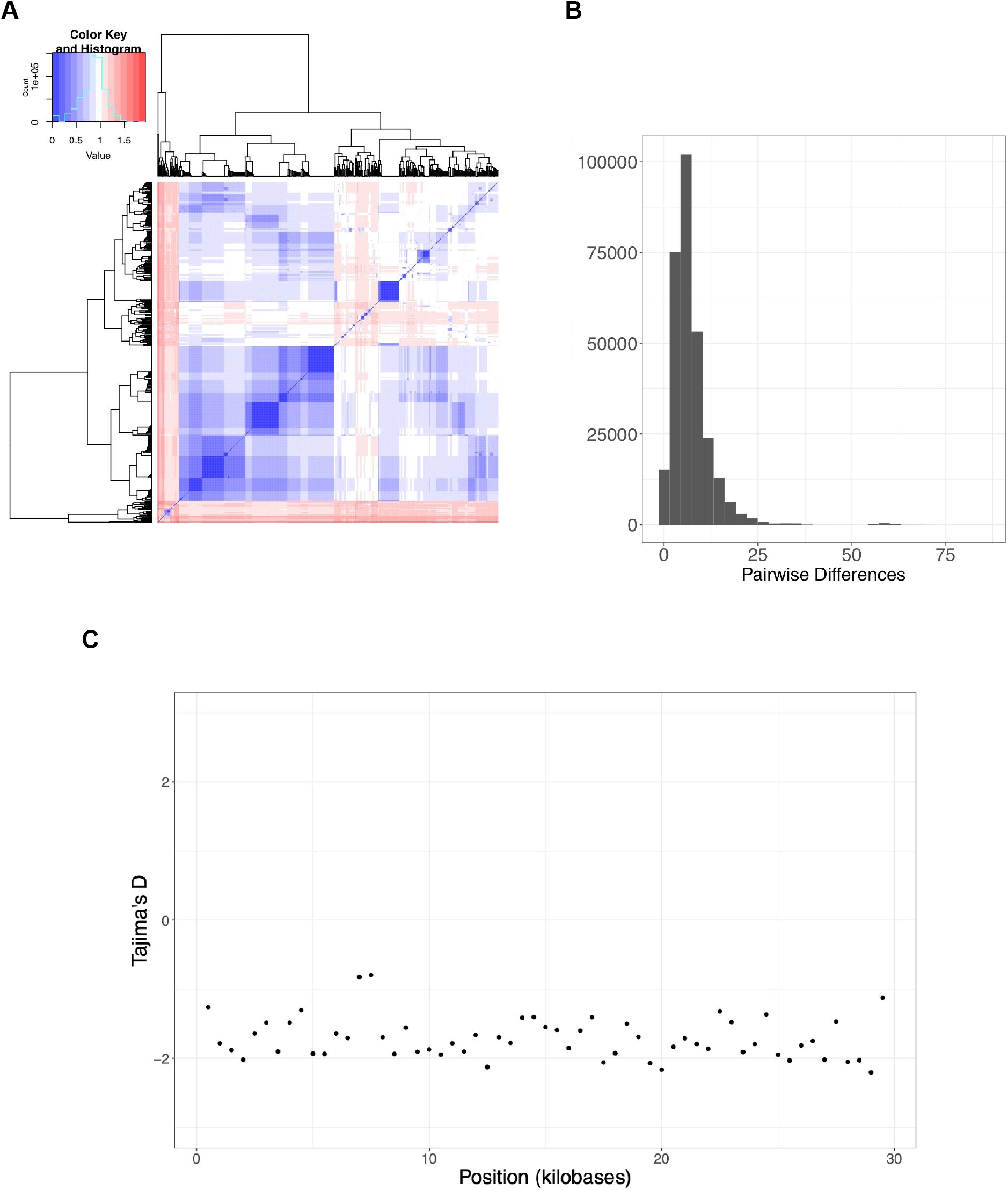
**A**. Distance matrix of pairwise distances for all complete genomes (>98% complete) from unique individuals in this study. **B**. Histogram of pairwise distances for all possible pairwise comparisons between complete genomes in the study. **C**. Tajimas’s D values in 500-base-pair intervals across the genome.

**Fig S5.**
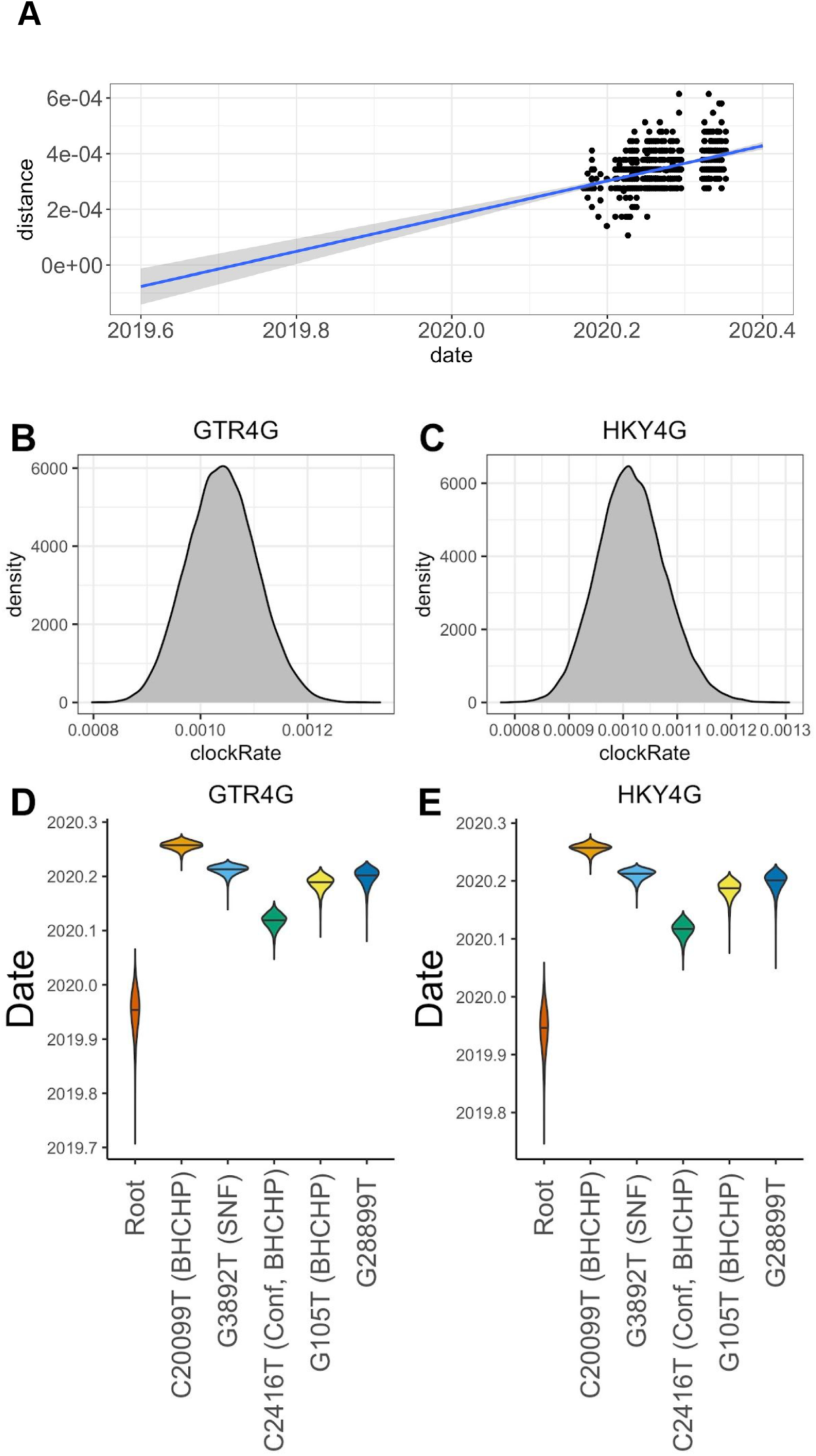
**A**. Linear regression of root-to-tip distance vs. date of sampling. Root-to-tip distance was calculated using TempEst (*44*) on maximum likelihood trees inferred using PhyML (*43*).

**Fig S6.**
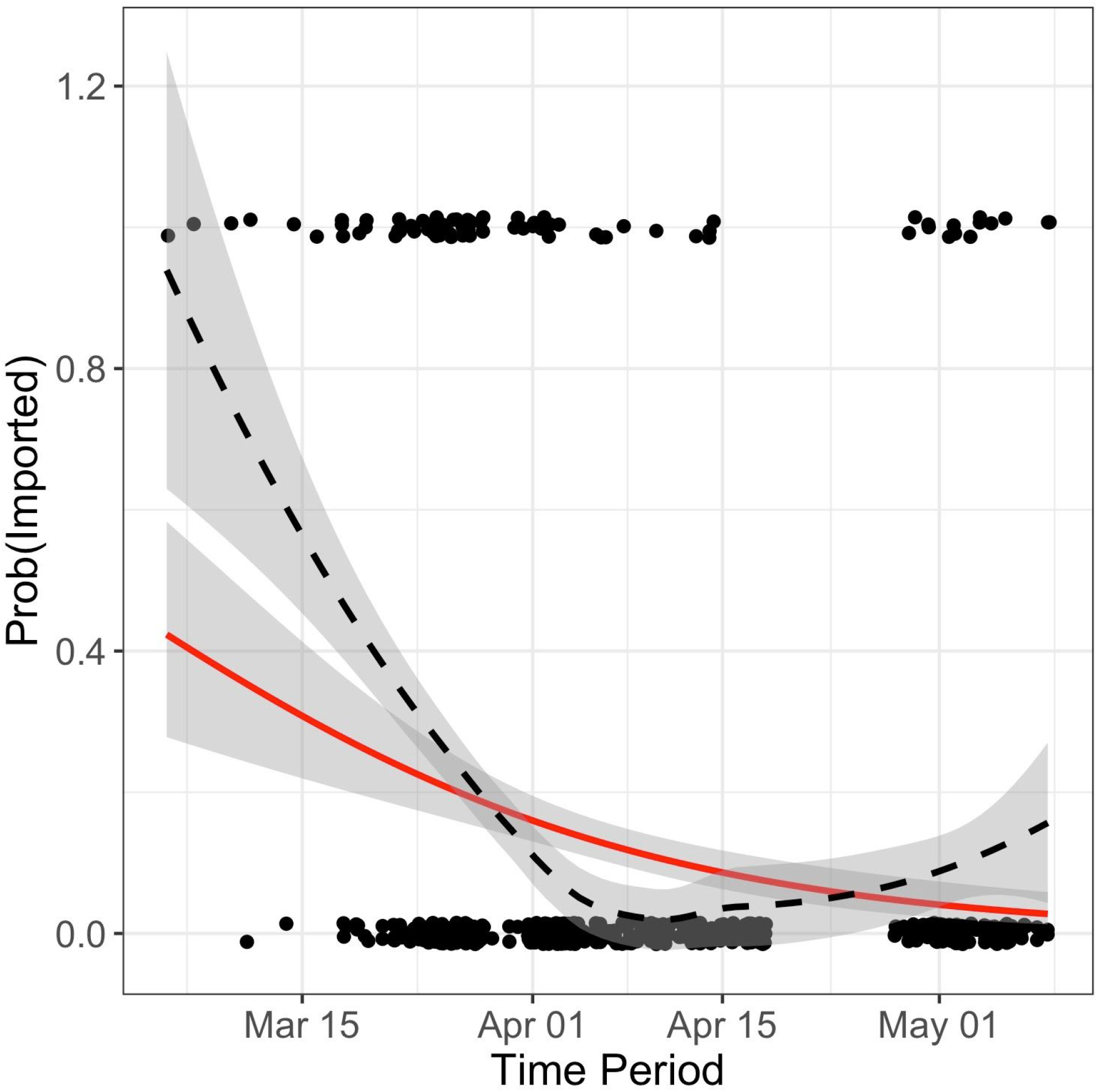
Probability of an importation event over time. Samples whose ancestral state was inferred as non-MA are coded as 1 and samples whose ancestral state is inferred as MA are coded as 0 (a small amount of noise is added to the y-coordinate to show the density of the data). A logistic regression (red curve) shows the probability of importation decreasing through the study period (*β*_1_.04999 +/- 0.01056, p = 2.2e-06). A loess smoother is shown with a dashed line.

**Fig S7.**
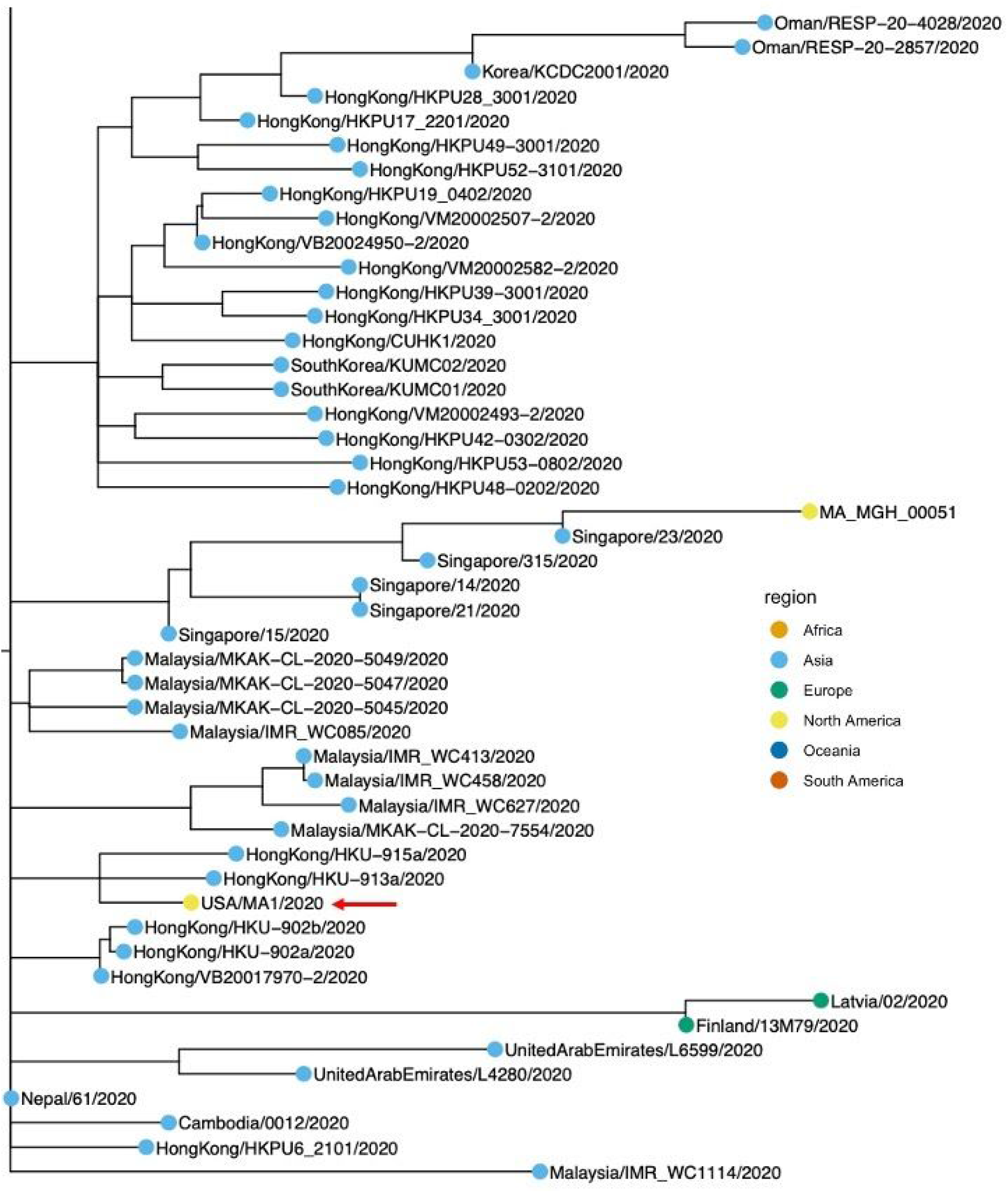
Portion of global time-stamped phylogeny (inferred using augur (*47*) with GISAID and MA genomes) containing MA-1 (red arrow).

**Fig S8.**
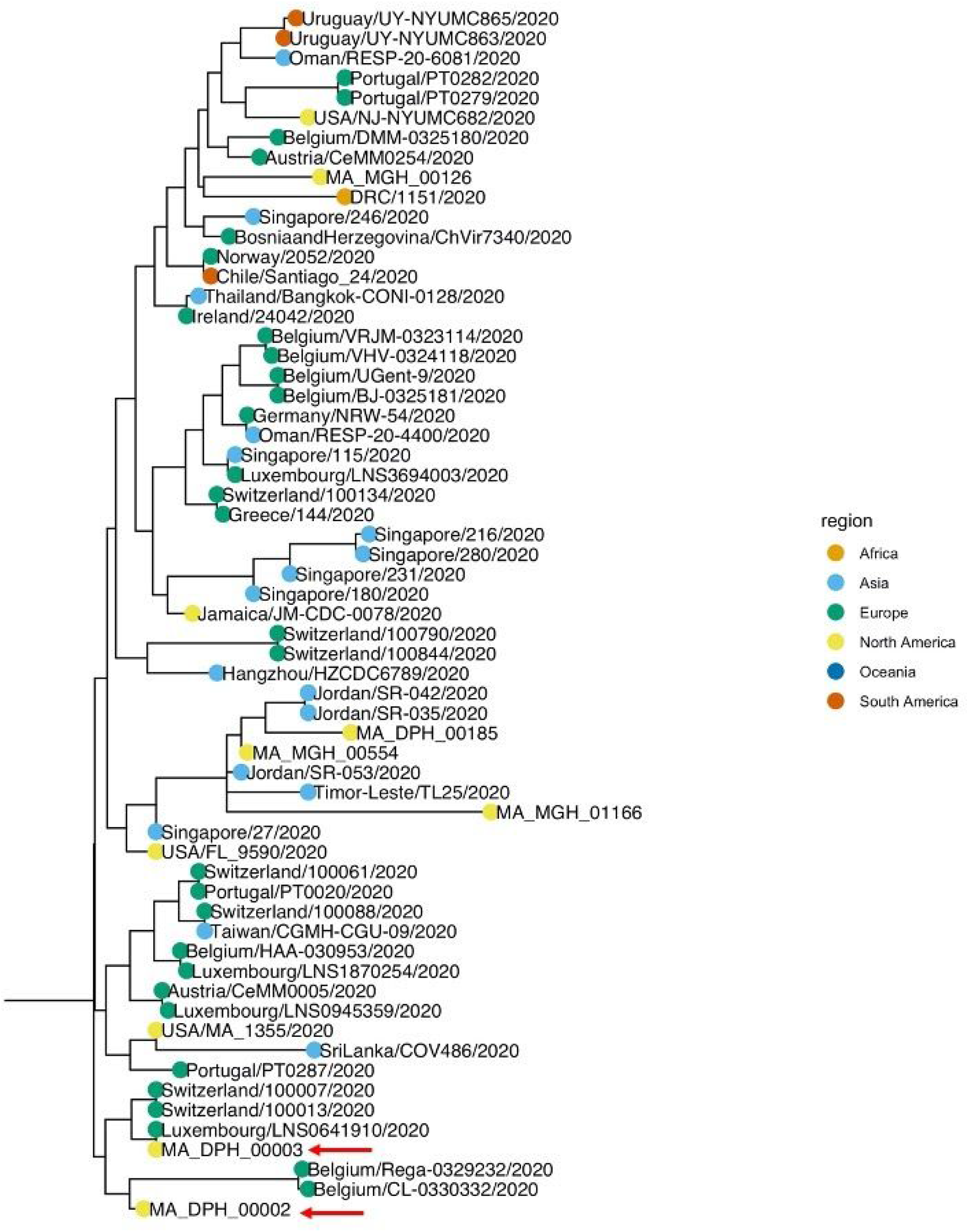
Portion of the global time-scaled phylogenetic tree (inferred using augur (*47*) with GISAID and MA genomes) containing DPH_00002 and DPH_00003 (marked with red arrows).

**Fig S9.**
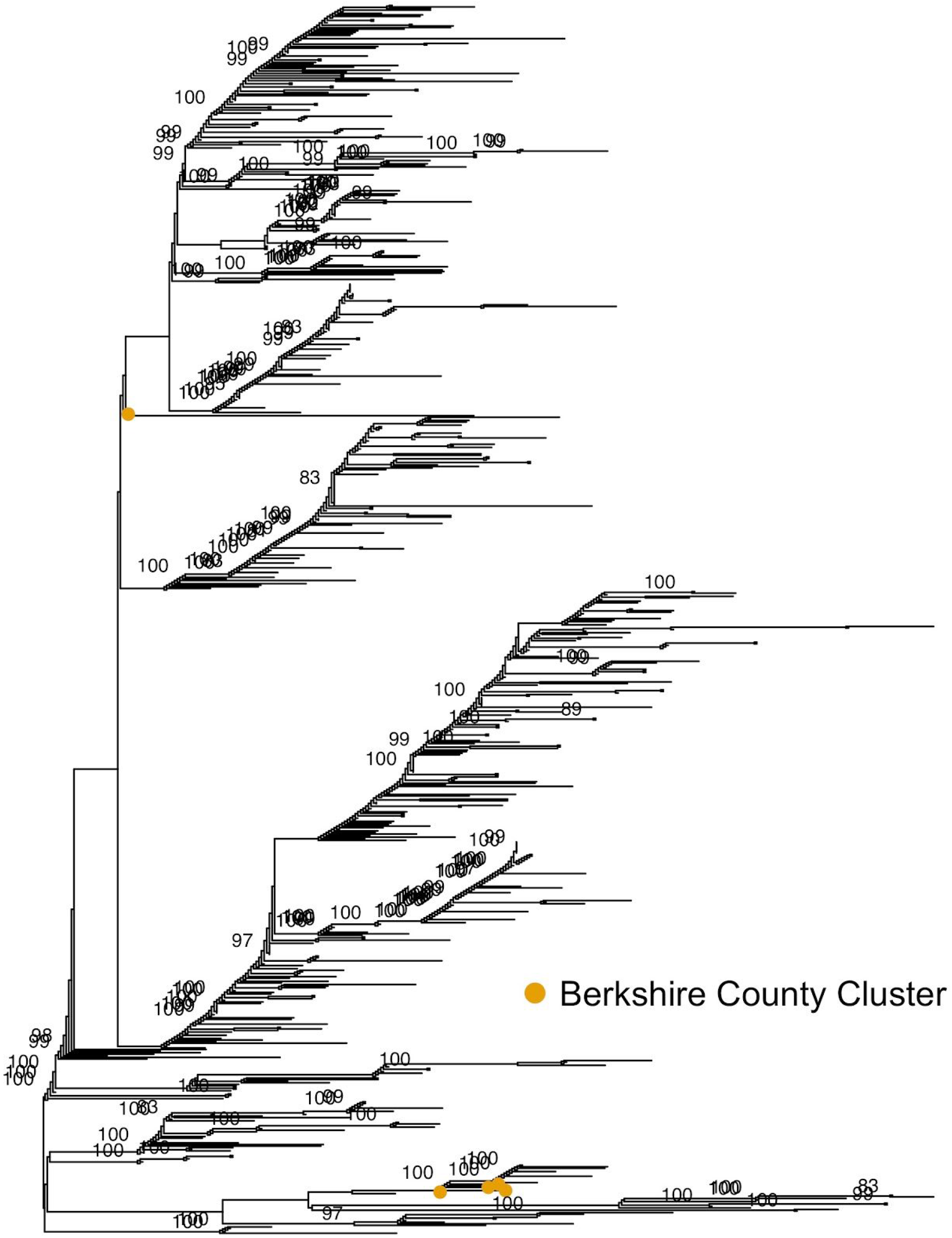
Phylogenetic ML tree of MA samples from this study plus partial genomes (>5kb) from Berkshire County Cluster. Ultrafast bootstrap support (when > 80) is shown at nodes. Tips corresponding to samples collected from the Berkshire County cluster are shown in orange.

**Fig S10.**
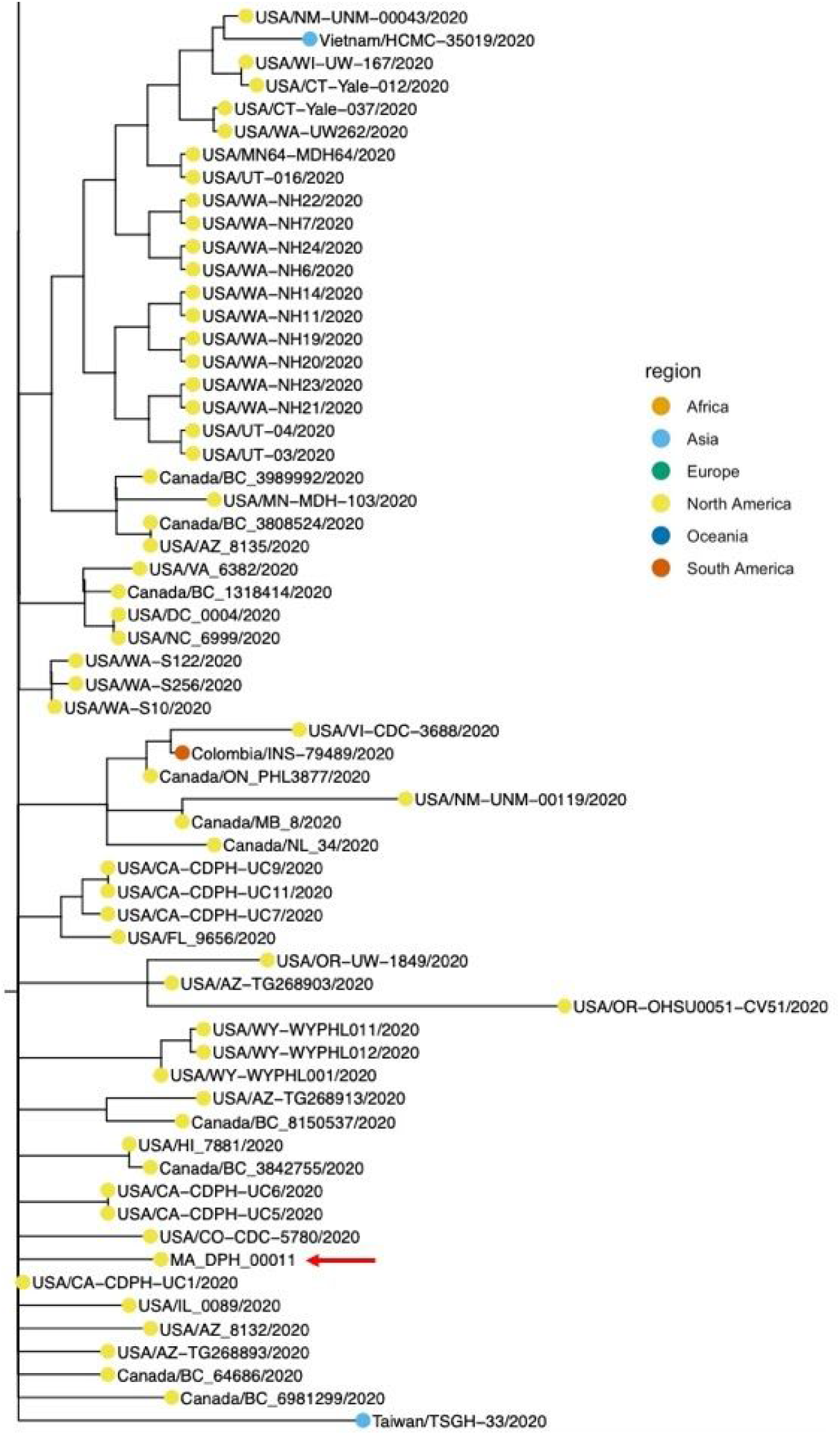
Phylogenetic position of DPH_00011 (type genome for the Western MA cluster, sample labeled with red arrow).

**Fig S11.**
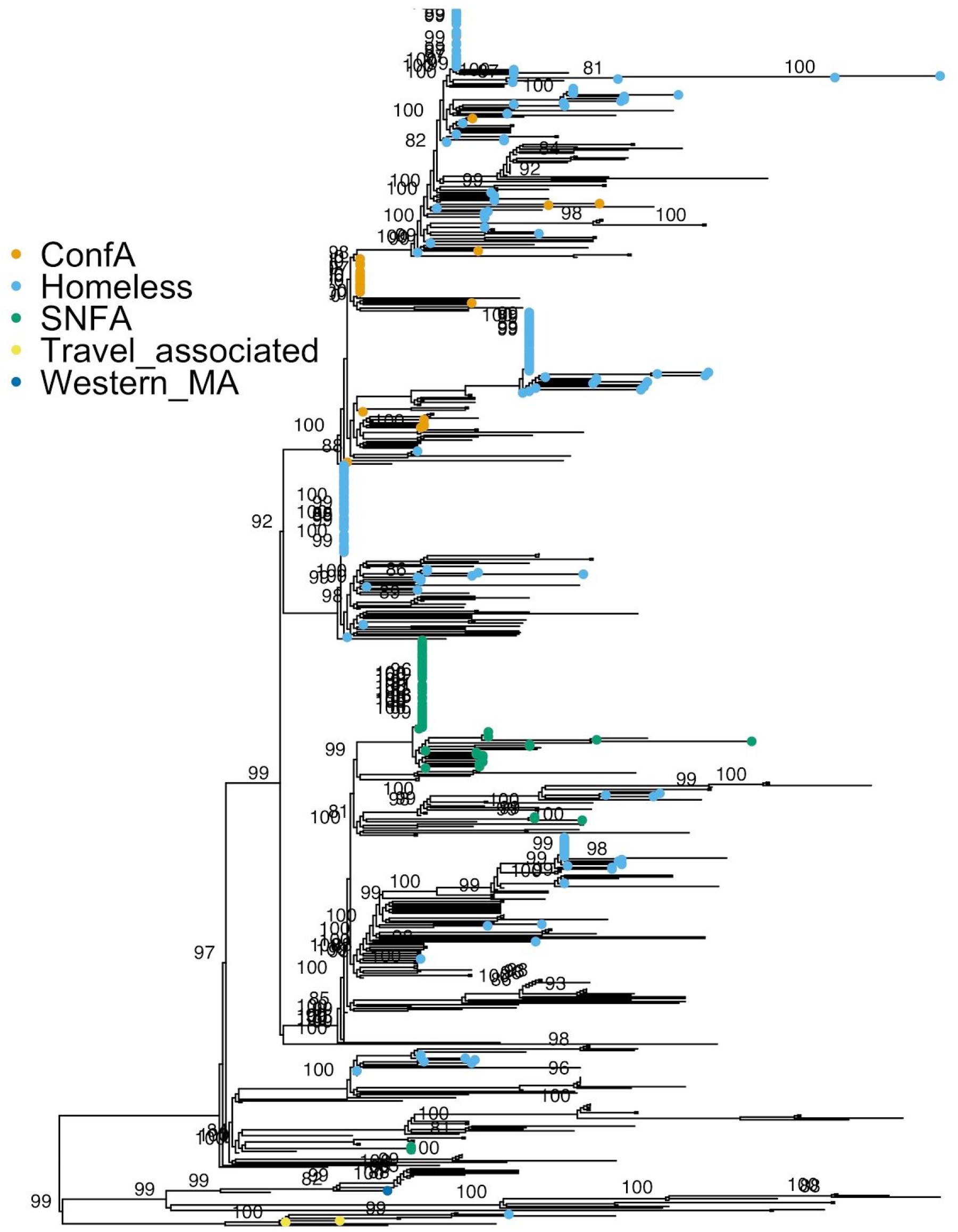
ML Tree of MA genomes computed using IQtree, with ultrafast bootstrap support shown at nodes with support > 80.

**Fig S12.**
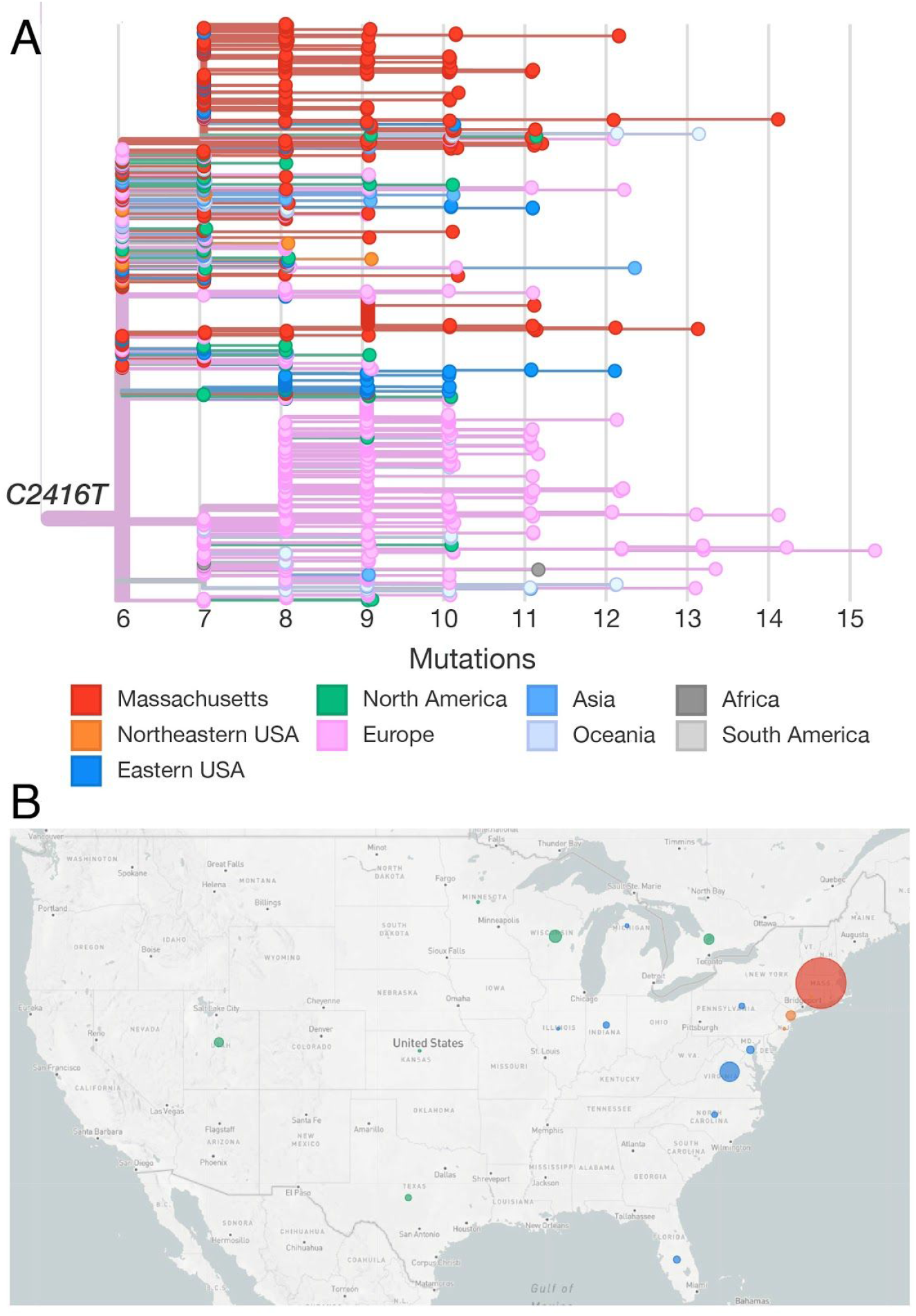
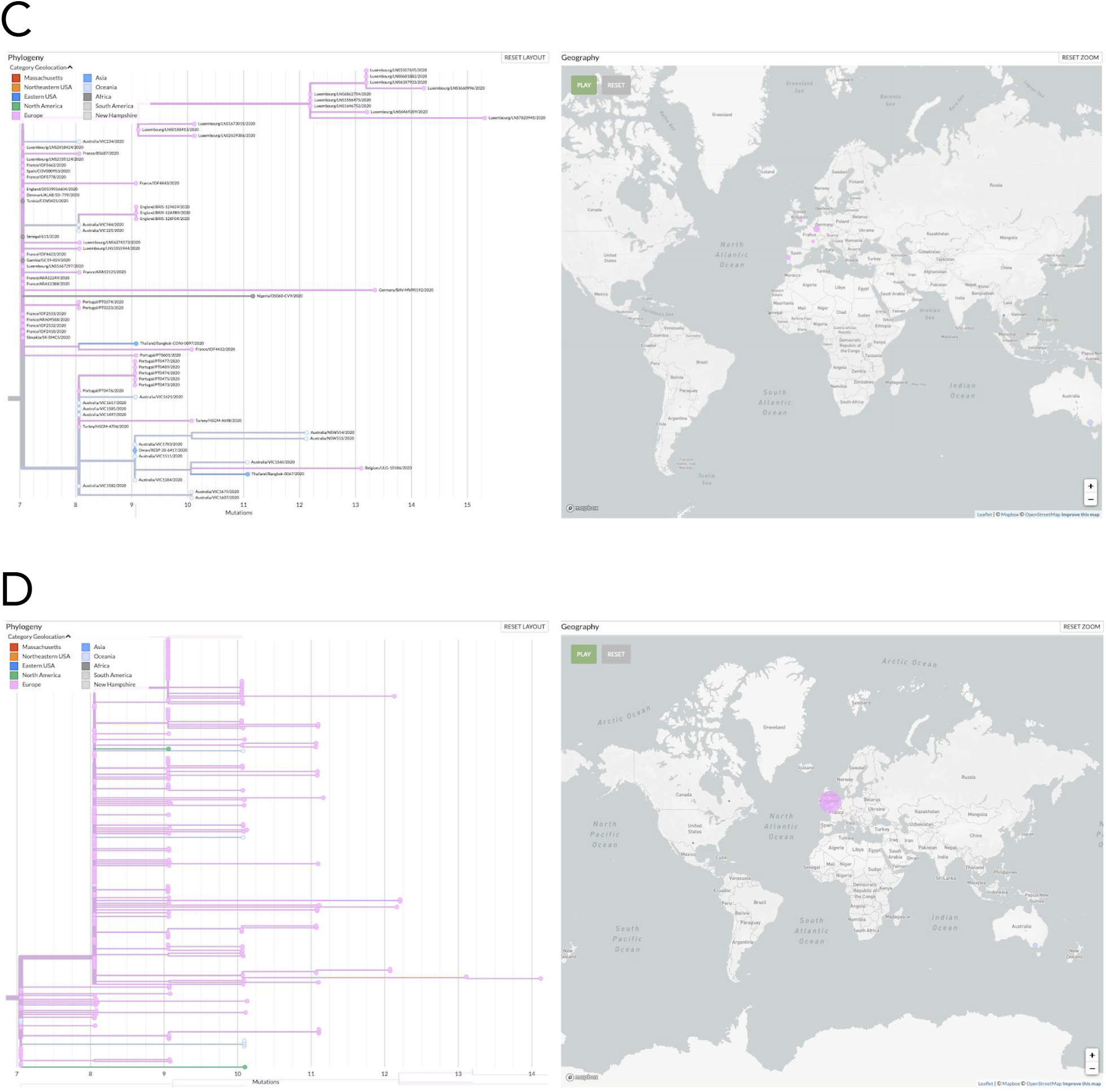
**A**. Divergence tree of the C2416T variant showing all global sequences (in GISAID through June 14, 2020) with the C2416T variant. **B**. Map showing the distribution of the C2416T variant across the United States. Circle size reflects the number of reported genomes per state. **C**. Phylogeny (left panel) and map showing global distribution of C2416T/G8371T. **D**. Phylogeny (left panel) and map showing global distribution of C2416T/G20578T.

**Fig S13.**
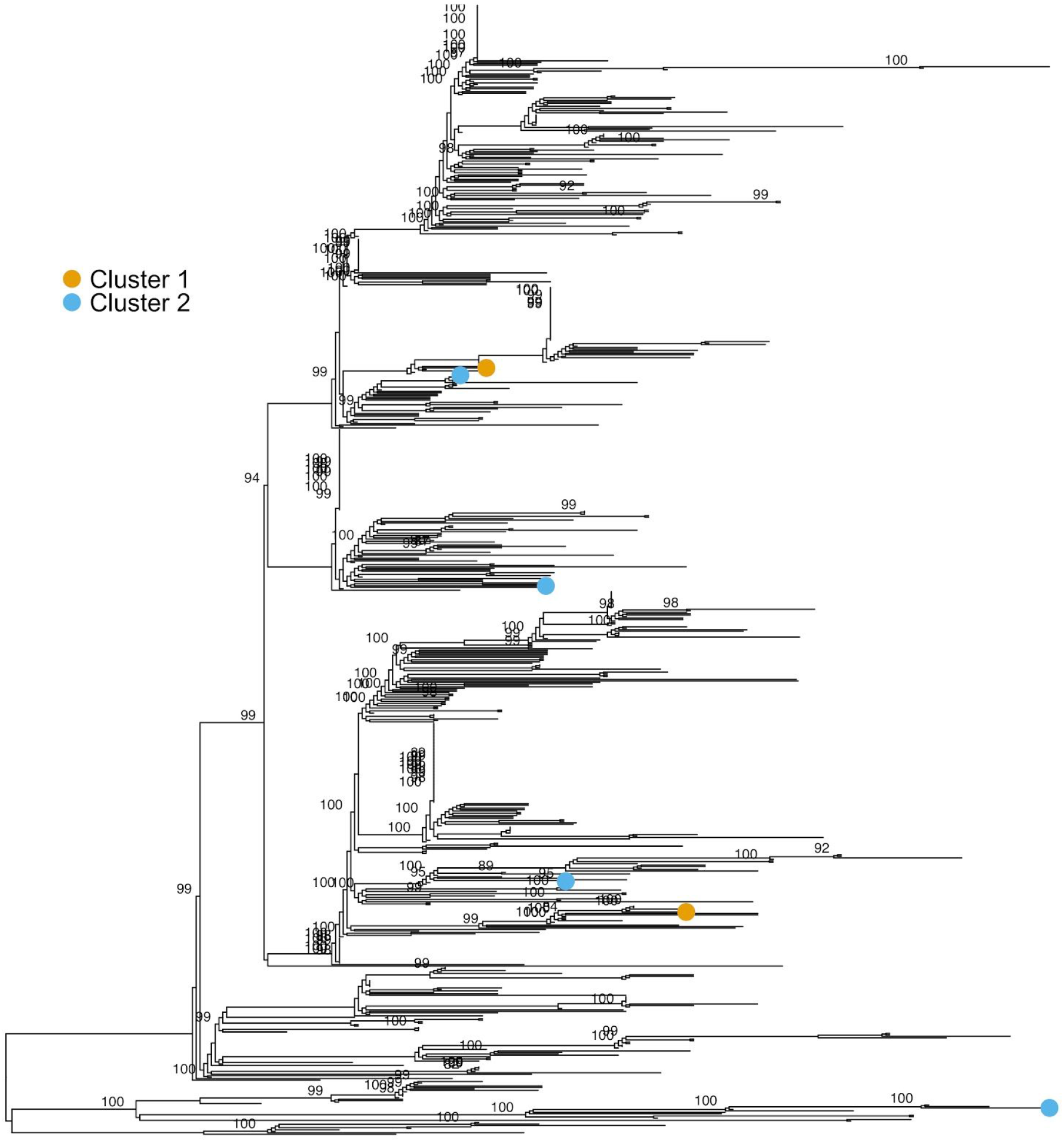
ML with samples from MGH suspected nosocomial clusters labeled. Posterior support for strongly supported (ultrafast bootstrap support > 80) nodes is shown.

**Fig S14.**
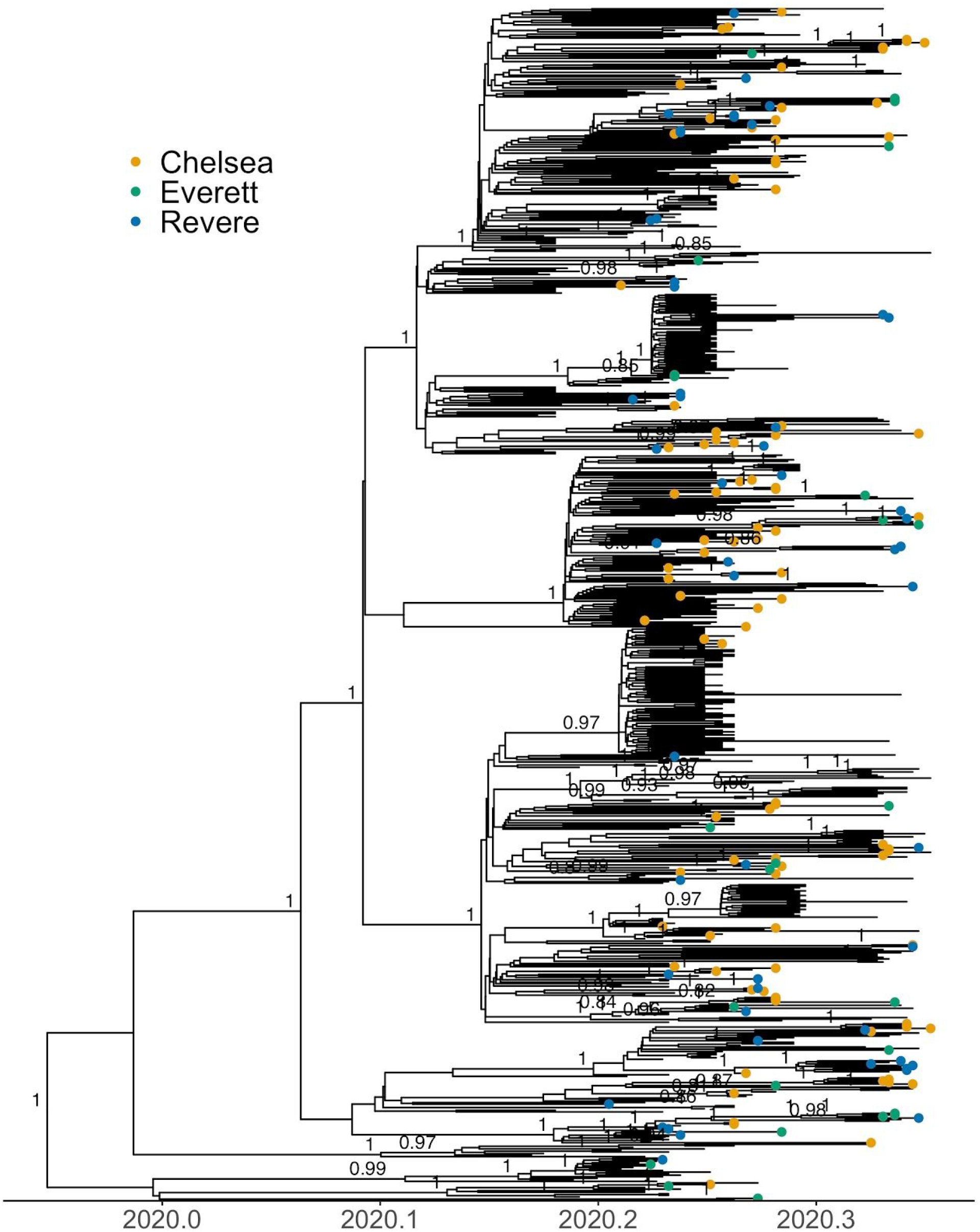
Sequenced samples labeled by zip code of residence for the top three zip codes in the set of 772 genomes from unique patients.

**Fig S15.**
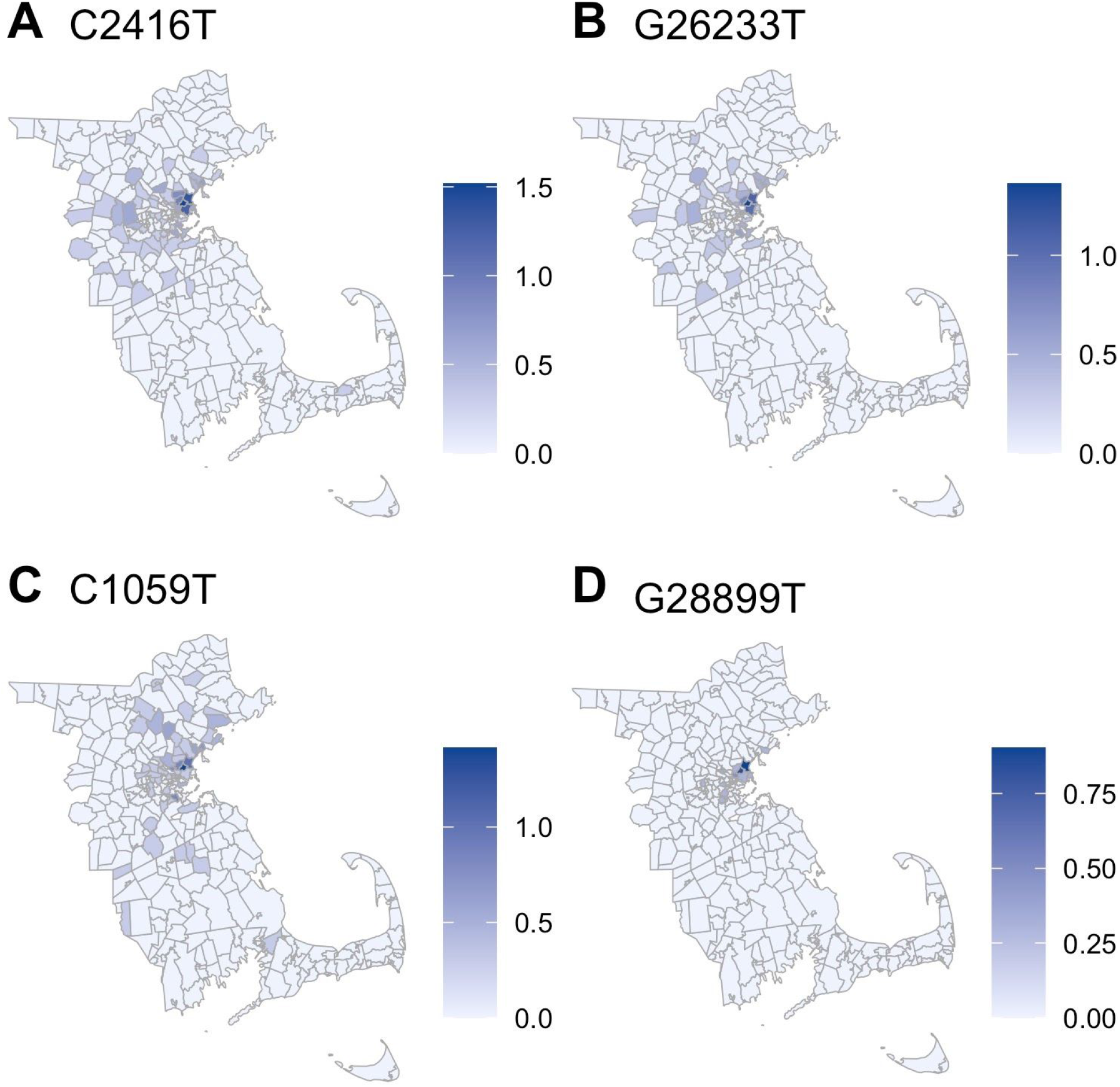
Geographic distribution of select lineage-defining variants in Eastern Massachusetts. The scale is in log10(case counts + 1).

**Fig S16.**
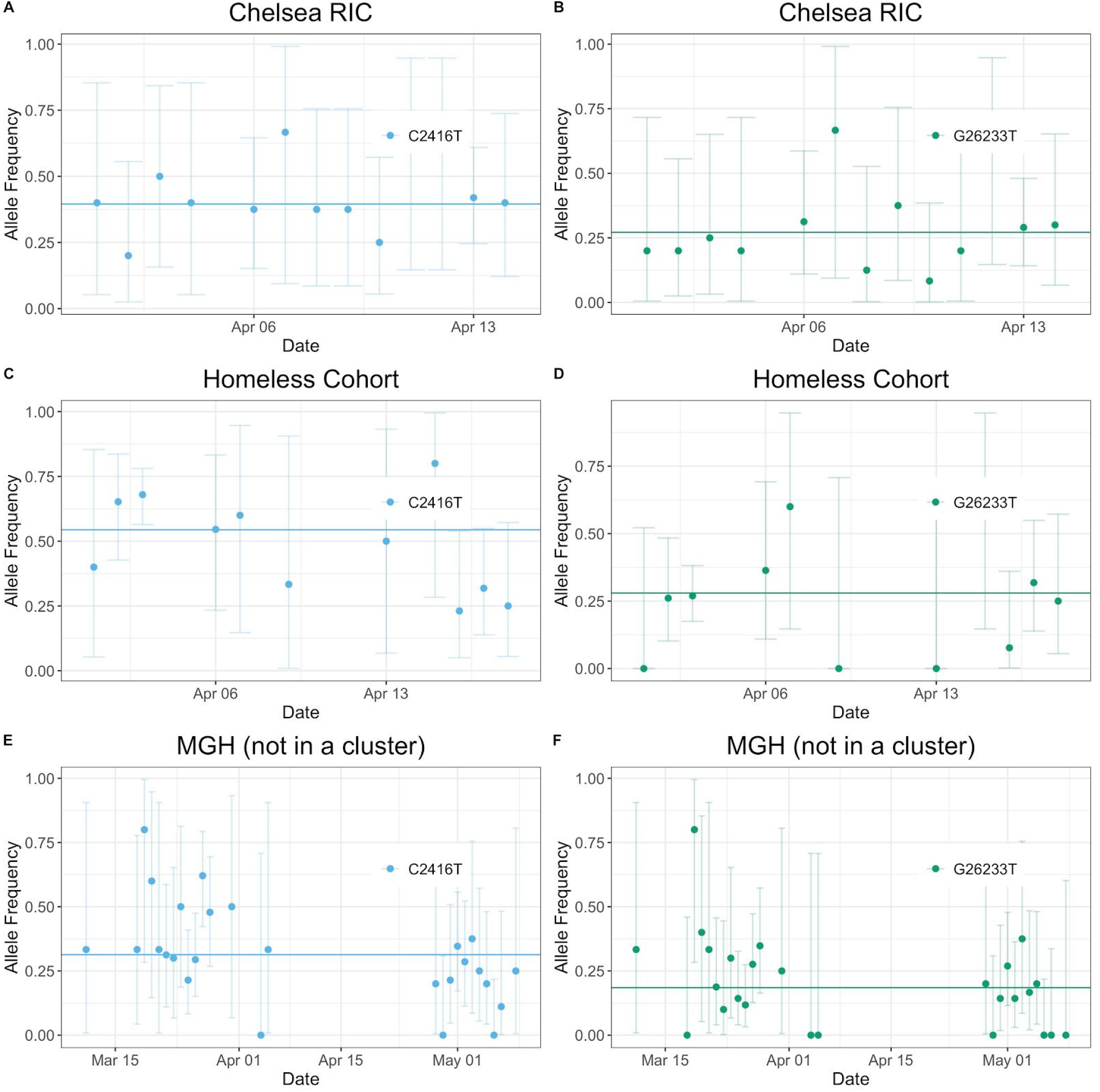
Prevalence of conference-associated variants by day among the Chelsea Respiratory Illness Clinic (RIC), among individuals experiencing homelessness sampled by BHCHP, and among samples available from the MGH Microbiology Laboratory that were not a part of known clusters (Conference, SNF) or from the Chelsea RIC. The solid line gives the cumulative allele frequency in each group.

**Fig S17.**
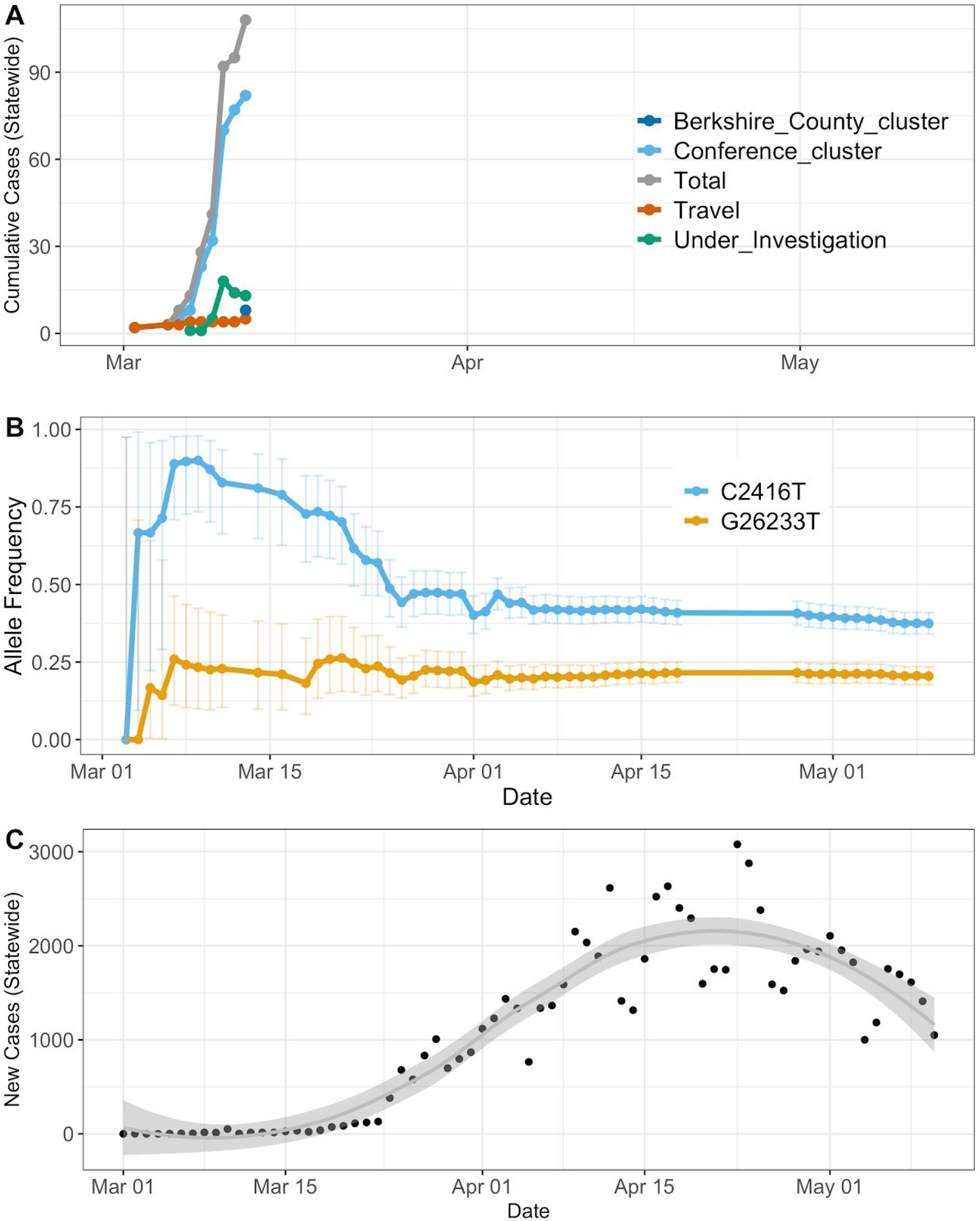
**A**. Cumulative case numbers by exposure group from March 9 through March 12 (period of data availability for the given exposures). **B**. Cumulative allele frequency of conference-associated alleles vs. time. **C**. Number of new infections reported by MADPH vs. time.

**Fig S18.**
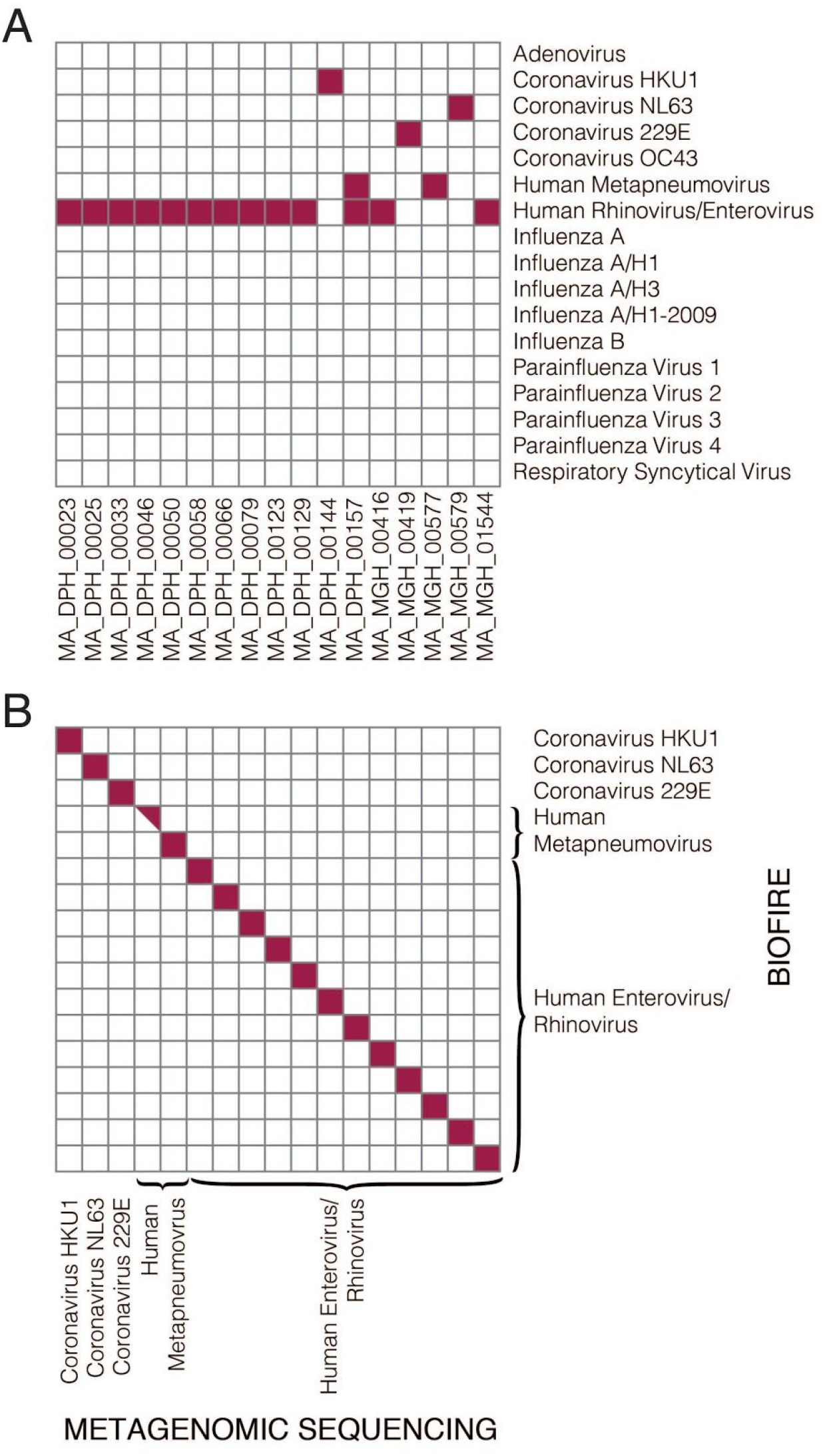
Confirmation of respiratory virus detection in metagenomic sequencing results. **A**. Results of the BioFire FilmArray Respiratory Virus Panel performed on the 17 available samples for which co-infections were detected by metagenomic sequencing. **B**. Concordance between BioFire and metagenomic sequencing results for respiratory viruses.

**Table S1**.

Download of sample_set table with summary assembly variables.

**Table S3:**

Table of geographic ancestral trait inferences.

**Table S3**.

Table of counts for all viral species classified by Kraken2.

